# United Kingdom National Register Study of Anti-Epileptic Medications: Suspected Foetal Congenital And Pregnancy-Associated Side Effects

**DOI:** 10.1101/2024.03.26.24304895

**Authors:** Benjamin Phillips, Ismay Evans, Victoria Skerrett, Alan M. Jones

## Abstract

**Objective:** There continue to be concerns regarding exposure during pregnancy to anti-epilepsy drugs (AEDs). The study aims were to determine the suspected adverse drug reactions (ADRs) associated with AEDs and potential mechanistic hypotheses.

**Methods:** Suspected ADR profiles for 8 AEDs were data-mined from the MHRA Yellow Card scheme (January 2018-August 2022) together with prescribing data from OpenPrescribing (August 2017-July 2022). The physicochemical, pharmacokinetic, and pharmacology of the AEDs were data-mined from public databases.

**Results:** The suspected ADRs per 1,000,000 *R_x_* identified across all AEDs are statistically significant (χ^2^ test, *P* < .05). Pregnancy, puerperium & perinatal conditions associated with lamotrigine (1.51 per 1,000,000 *R_x_,* χ^2^ test, *P* < .05, *d* = 2.720, 95% CI [1.656, 4.469]) had a larger size effect than valproic acid (2.28 per 1,000,000 *R_x_,* χ^2^ test, *P* < .05, *d* = 1.846, 95% CI [1.150, 2.964]). The large size effect associated with valproic acid for congenital and hereditary disorders (*d* = 9.069, 95% CI [5.807, 14.163]) and foetal exposure during pregnancy (*d* = 6.632, 95% CI [4.894, 8.988]) were notable amongst the AEDs. Valproic acid, a known teratogen, had the unique and clinically achievable targeting of histone deacetylase (HDAC 1 IC_50_ = 54.4, HDAC2 IC_50_ = 82.4 micromolar, HDAC3 IC_50_ = 148 micromolar, HDAC8 IC_50_ = 144 micromolar, C_max_ = 184.3 micromolar) associated with teratogenicity.

**Significance:** There is renewed discussion about the management of epilepsy in pregnancy, and the risks of different AEDs. Whilst 1 in 250 women have epilepsy, they account for 1 in 10 of women who die in childbirth or postpartum. Fears about ADRs impact on adherence to medication, whilst pregnancy itself reduces the serum level of AEDs. As a result of this women are at increased risk of seizures during pregnancy and childbirth. There has been a doubling of Sudden and Unexpected Death in Epilepsy (SUDEP) in mothers between 2013-2015 and 2019-2021 in the UK and Ireland. The AEDs studied have diverse modes of action, and the unique polypharmacology of AEDs influences their ADR profiles. Lamotrigine had a larger size effect than valproic acid (*d* =2.720 vs 1.846) for suspected pregnancy, puerperium and perinatal ADRs. As noted in other studies, there is a suspected association between valproic acid exposure and 1) congenital and hereditary disorders (*d* = 9.069), and 2) foetal exposure during pregnancy (*d* = 6.632) compared to other studied AEDs. Pregnancy-related ADRs with levetiracetam and topiramate did not reach statistical significance, however neurological ADRs in children who were exposed to lamotrigine and levetiracetam continue to be the subject of scrutiny.

**Key Points:**

1. There are ongoing concerns regarding exposure to all anti-epilepsy drugs (AEDs) during pregnancy. Poor seizure control in pregnancy is a cause of maternal death, valproic acid continues to be used by women despite it being a known teratogen, and other AEDs also carry risks of significant ADRs.
2. AEDs have diverse modes of action, and the unique polypharmacology of AEDs influences their ADR profiles.
3. Lamotrigine had a larger size effect than valproic acid (*d* =2.720 vs 1.846) for suspected pregnancy, puerperium and perinatal ADRs.
4. There is a strong association between valproic acid exposure and congenital and hereditary disorders including foetal valproate spectrum disorder (FVSD), autism spectrum disorder (ASD), spina bifida, polydactyly and cleft palate (*d* = 9.069).
5. There is an association between valproic acid and foetal exposure during pregnancy (*d* = 6.632).

## Introduction

Adverse drug reactions (ADRs) are unintended responses to a drug, causing 1 in 6 hospital admissions, at a cost to the NHS of approx. £2.2 Bn p.a., increasing duration of hospital stays, contributing to poor adherence, and adversely affecting quality of life.[1]

Epilepsy affects 1 in 103 people globally.[2] Anti-epileptic drugs (AEDs) control seizure activity in 70% of patients but can also cause adverse drug reactions (ADRs).[2] AEDs prescribed in the United Kingdom include valproic acid, carbamazepine, gabapentin, lamotrigine, levetiracetam, oxcarbazepine, topiramate and zonisamide (**Figure S1**).

Over 2,500 women who suffer from epilepsy become pregnant each year in the UK [3] and the prescribing of AEDs in pregnancy is a cause for concern as many of them have teratogenic effects. *In-utero* AED exposure increases the risk of a physical birth abnormality, harms growth and development.[3] Women with epilepsy are ten times more likely to die during pregnancy and the risks to the developing foetus need to be balanced against the risk to both mother and baby of severe seizures.[3–4]

Sodium valproate is a known teratogen, and a drug of concern for many decades, being first linked to increased risks of congenital and developmental disorders in 1972.[5]. Subsequent studies led to teratogenic risk warnings to be added to valproic acid.[6] Currently 1,800 women of child-bearing age take valproic acid despite the Pregnancy Prevent Programme (PPP) being in place since 2018.[7] I*n-utero* exposure can cause foetal valproate syndrome, which increases the risk of congenital and neurodevelopmental disorders, and approximately 20,000 babies in the UK have been affected.[8] 40 in 100 babies exposed to valproic acid *in-utero* have neurodevelopmental disorders.[9–10] Valproic acid has the highest risk of congenital malformations, including spina bifida, affecting over 10 in 100 babies.[8] Valproic acid’s *in-utero* effects are dose dependent, with higher doses increasing risks of congenital or developmental disorders.[11]

The prescription of valproic acid to women of childbearing potential is now accompanied by a Pregnancy Prevention Programme (PPP). [12–14] Through this, the use of sodium valproate in pregnancy has seen a significant decline;[15] however, it is still used in pregnancy. Despite a high-profile safety review,[16] medication warnings and messages to professionals, many pregnant women with epilepsy have not had the opportunity to discuss valproate with a specialist. A survey by three UK epilepsy charities found that nearly 20% of women prescribed valproate reported they were not aware of the risks of the drug, and that only 41% had signed up to the PPP.[15]

Valproic acid is the most effective medication for tonic-clonic seizures[17] which impacts on prescribing choices. Focal epilepsy can be effectively managed with alternative AEDs, however there are concerns about the ADR profiles of alternatives to valproic acid. Lamotrigine and levetiracetam are considered the safest AEDs.[3] However, levetiracetam and lamotrigine are drugs named for additional scrutiny for neurodevelopmental ADRs in children exposed in utero in the EMPiRE project [18] Topiramate and carbamazepine are high risk and cause physical defects in 4-5 per 100 babies compared to 2-3 in 100 in the general population.[3] Bjørk MH, et al[19] showed topiramate and valproic acid increase risks of autism spectrum disorder (ASD) and neurodevelopmental disorders, initiating a safety alert review into topiramate in the UK.

Belete et al.[20] found evidence of an association between AED exposure (carbamazepine, lamotrigine, levetiracetam, and valproic acid) and incident Parkinson disease using data from the UK Biobank. This wide range of ADRs across all the AEDs, with very significant consequences for mothers and their children, suggests a combinatorial data-driven assessment of AED polypharmacology and ADR profiles [21–26], is timely.

## Methods

### Prescribing Data

NHS (England) primary care prescribing data (August 2017-July 2022 inclusive) for the generic product form was obtained from the OpenPrescribing database https://openprescribing.net/ on 13.10.2022.

### Suspected Adverse Drug Reactions

Suspected ADR data was extracted from the MHRA Yellow Card database https://yellowcard.mhra.gov.uk/idaps (January 2018-August 2022 inclusive on 15.10.2022). A lag time of 5-months between reaction and report was factored into the data. AEDs that did not meet the inclusion criteria (**Table S1**) were removed (**Table S2**).

Data were collected for the number of suspected ADRs and stratified by organ class and fatalities. Significant suspected ADRs were found by MedDRA organ class with > 100 total reactions in ≥ 1 AED studied or statistical significance (*p* < .05, χ^2^) across the AEDs studied. Pregnancy was included despite low suspected ADR reports and did not show statistical significance (*P* = .655, χ^2^) for an important research comparison.

ADR Data were standardised per 1,000,000 prescriptions to enable comparisons uninfluenced by prescribing variation (**Equation S1**).

### Chemical Properties

#### Physiochemical and pharmacokinetic properties

AED properties were determined from the summary of product characteristics (SPC) accessed through the Electronic Medicines Compendium (EMC) and the manually curated database of bioactive molecules with drug-like properties of the European Molecular Biology Laboratory (ChEMBL) https://www.ebi.ac.uk/chembl/.

To calculate parameters:

The –log_10_ of the half-maximal inhibitory concentration (*p*IC_50_) was gathered for the main human target protein of each AED using the formula −*log*_10_(*IC*_50_).

Lipophilic Ligand Efficacy (LLE) was found through *p*IC_50_ – clog_10_P, with cLog_10_P the calculated partition coefficient between lipid (*n*-octanol) and the aqueous phase. LLE values (<5) correlate to increased off-target drug-protein interactions and an increased risk of toxicity.[27]

Peak serum concentration of the drug (C_max_) values was gathered from either the SPC and/or FDA monographs and converted to nM.

### Target Affinity

Each AED’s target affinity was obtained *via* the IC_50_ between the AED and single human protein target from the ChEMBL database and relevant literature (accessed 06.11.2022). AEDs with multiple IC_50_ values for the same target were standardised to the median to give a relative affinity and compensate for outliers.

### Statistical Analysis

Chi-Squared tests (χ^2^) were performed on standardised ADR data to find statistical significance (*p* < .05, χ^2^) using Microsoft Excel. Where Pearsons χ^2^ tests were performed on *n* < 5, Yates’s correction was employed to avoid the assumption of continuous distribution of binary data.

Odds Ratio (*d*) and confidence intervals [CI] were calculated using **Equation S2**. This method was employed to find within the 8 AEDs which, if any, have a significantly higher risk of a particular ADR compared to other AEDs. An alternative approach to detect a “true” signal by comparison of a particular ADR against all UK-licensed drugs was outside the scope of this study. It should also be noted that **Table S3** contains >2000 two-way comparisons between pairs of AEDs, of which > 300 have *P* < .05. Purely by chance ∼100 of these pairs would have a *P* < .05. Thus, additional odds risk ratio (*d*) and confidence interval (CI) statistical evaluation was performed on the most significant findings to determine CI >1 not ≥ 1 to avoid bias. Calculations were completed using Microsoft Excel and displayed as odds-ratio plots.

### Ethics Approval

All data used in this study was publicly available, with no identifiable patient information, and was exempt from ethics approval. Prepublication of a protocol for this study was not required based on the above ethical considerations.

## Results

### Physiochemical and Pharmacokinetic Properties

**Table S4** holds a summary of the main AED properties and gives a reference dataset to the drug concentrations that would have been likely to have affected the targets studied.

### Target Affinity

It should be noted that the C_max_, measured as total (bound and unbound) plasma concentration (usually after a single dose), and primarily related to the volume of distribution of total drug, is a good surrogate for the tissue concentrations likely to be achieved during short-term therapy. However, during long-term therapy, and factoring in interactions on the pharmacological targets, the C_max_ is likely to drift higher due to bioaccumulation.

Valproic acid’s main target is histone deacetylases (HDAC) 1-9 (**Table 1**) each HDAC isozyme responsible for different developmental disorders.[28]

**Table 1.**
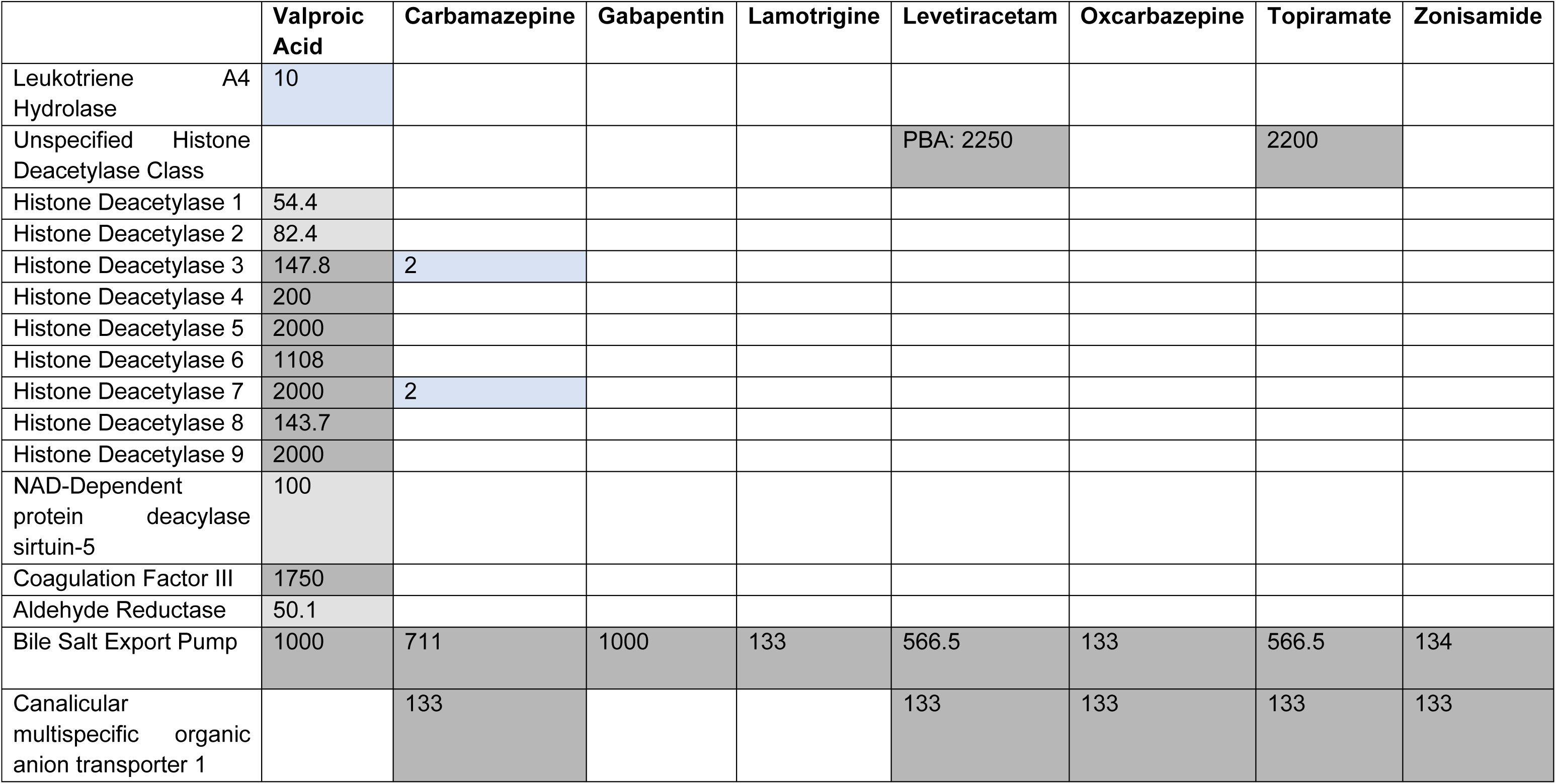
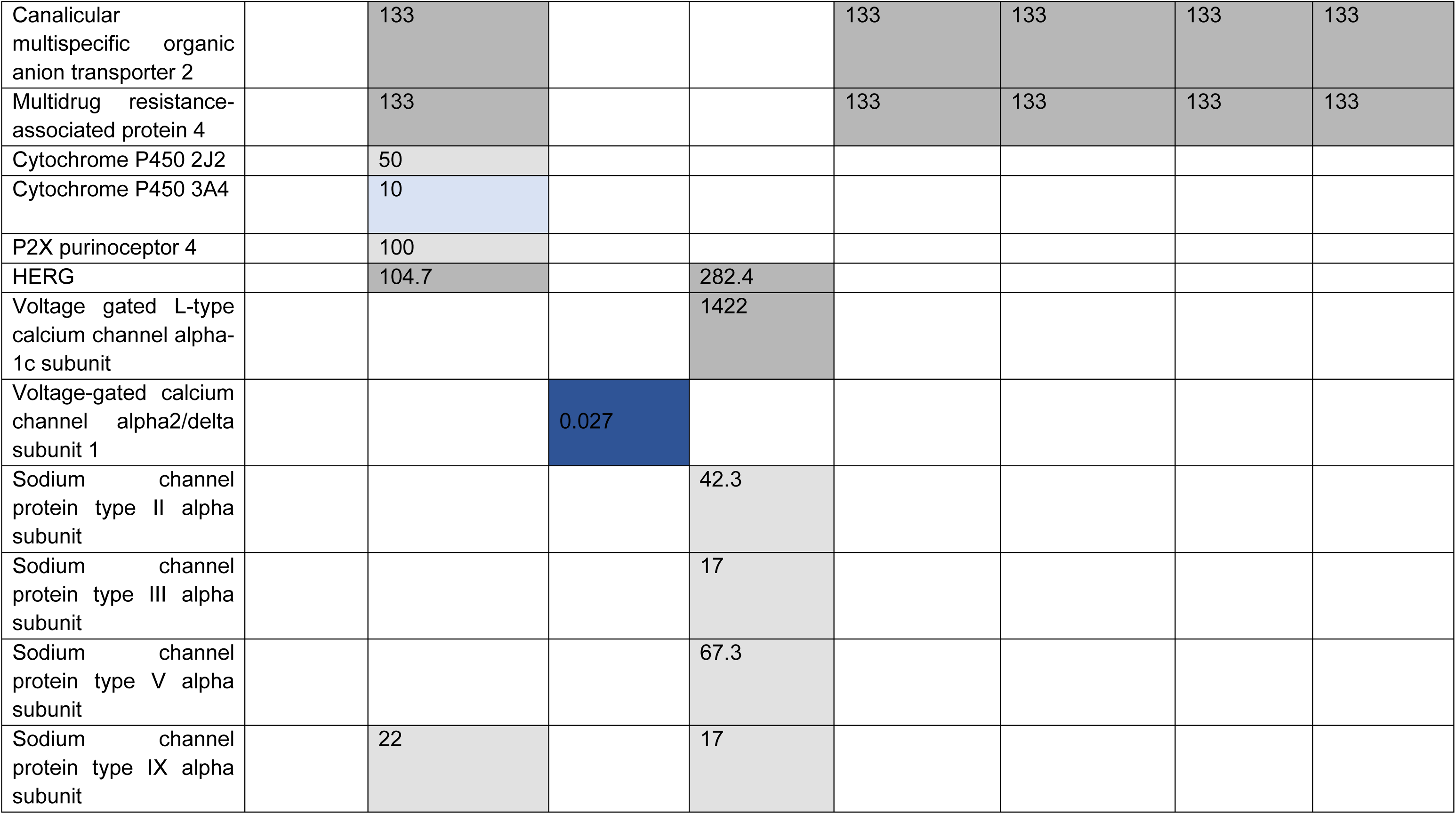
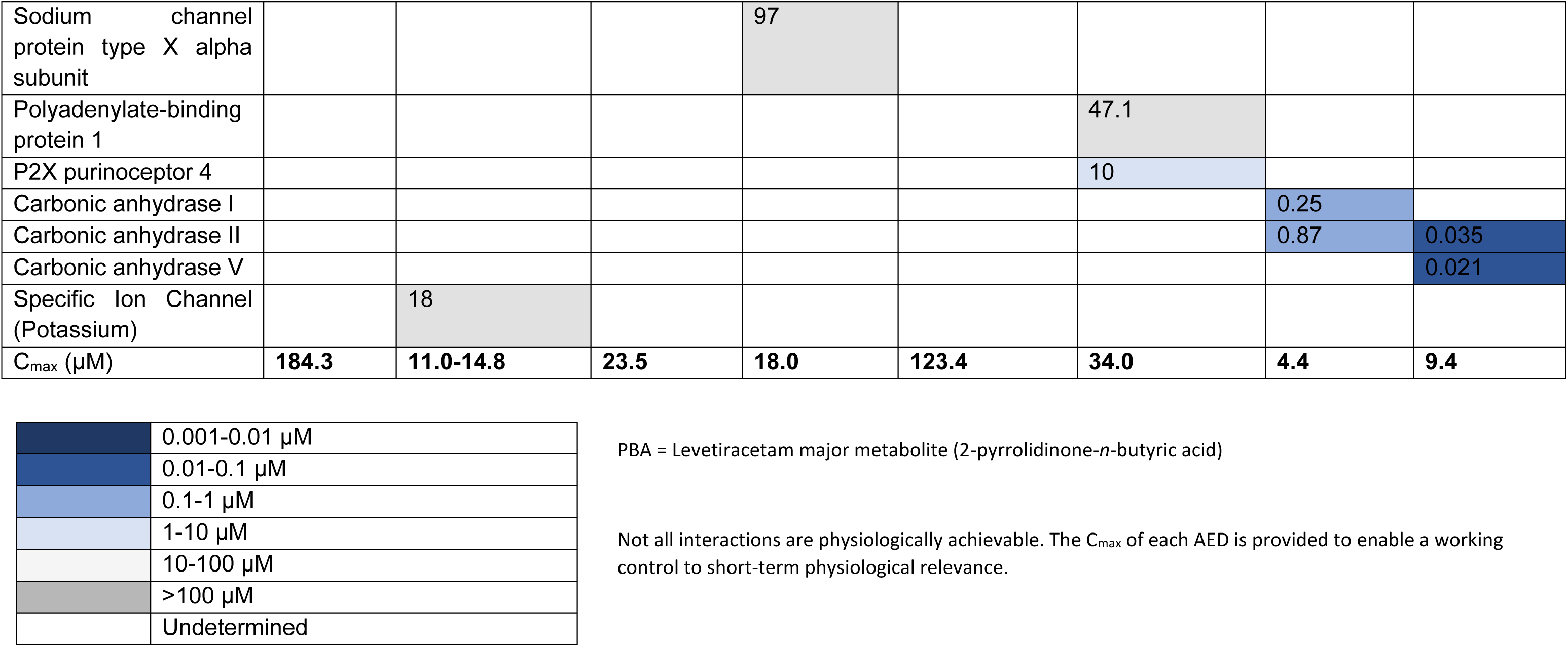
Selectivity profiles of the 8 AED’s based on human single protein targets and standardised as median IC_50_ values (µM)

Valproic acid teratogenic effect has been well known since 2001 [29–32] A cumulative meta-analysis of clinical trials by Tanoshima et al provides further confirmatory evidence of the link between valproic acid and teratogenicity.[33]

Valproic acid (HDAC1 IC_50_ = 54.4 µM, HDAC2 IC_50_ = 82.4 µM, HDAC3 IC_50_ = 147.8 µM, HDAC4 IC_50_ = 200 µM, HDAC5 IC_50_ = 2,000 µM, HDAC6 IC_50_ = 1,108 µM, HDAC7 IC_50_ = 2,000 µM, HDAC8 IC_50_ = 143.7 µM, and HDAC9 IC_50_ = 2,000 µM), carbamazepine (HDAC3 IC_50_ = 2 µM, HDAC7 IC_50_ = 2 µM), topiramate (unspecified HDAC IC_50_ = 2,200 µM), and levetiracetam’s active metabolite, 2-pyrrolidinone-*n*-butyic acid [PBA] (unspecified HDAC 2,25 µM) inhibit HDAC family members implicated in foetal defects. Only carbamazepine’s inhibition of HDAC3 and 7 (C_max_ = 11-14.8 µM) and valproic acid’s inhibition of HDAC1, 2, 3, and 8 (C_max_ = 184.3 µM) are clinically achievable.

All AEDs displayed unique pharmacological profiles. Valproic acid was the least selective, with fourteen protein target interactions compared to gabapentin’s two. Levetiracetam displayed no biologically relevant inhibition at any off target. All AEDs interact with bile salt export pump (BSEP), but no clinically significant inhibition occurs. Valproic acid (C_max_ = 184.3 µM) also had biological achievable inhibition of Leukotriene A4 Hydrolase (IC_50_ = 10 µM) and NAD-dependent protein deacylase sirtuin (IC_50_ = 100 µM).

## ADRs

### Total ADRs

Gabapentin is the most prescribed AED (36,692,340 *R_x_*) (**Table 2**). Oxcarbazepine had the most reported ADRs per million *R_x_* (393.90), zonisamide (248.43), valproic acid (247.86), topiramate (206.43), carbamazepine (162.87), levetiracetam (136.33), lamotrigine (115.14) and gabapentin (62.44), respectively. Zonisamide had the highest suspected fatality rate (3.57 per 1,000,000 *R_x_*) in contrast to oxcarbazepine (0). All AEDs had at least one organ system ADR which were more likely to occur with its use. Overall suspected ADRs per 1,000,000 *R_x_* identified across all AEDs are statistically significant (χ^2^ test, *P* < .05).

**Table 2.**
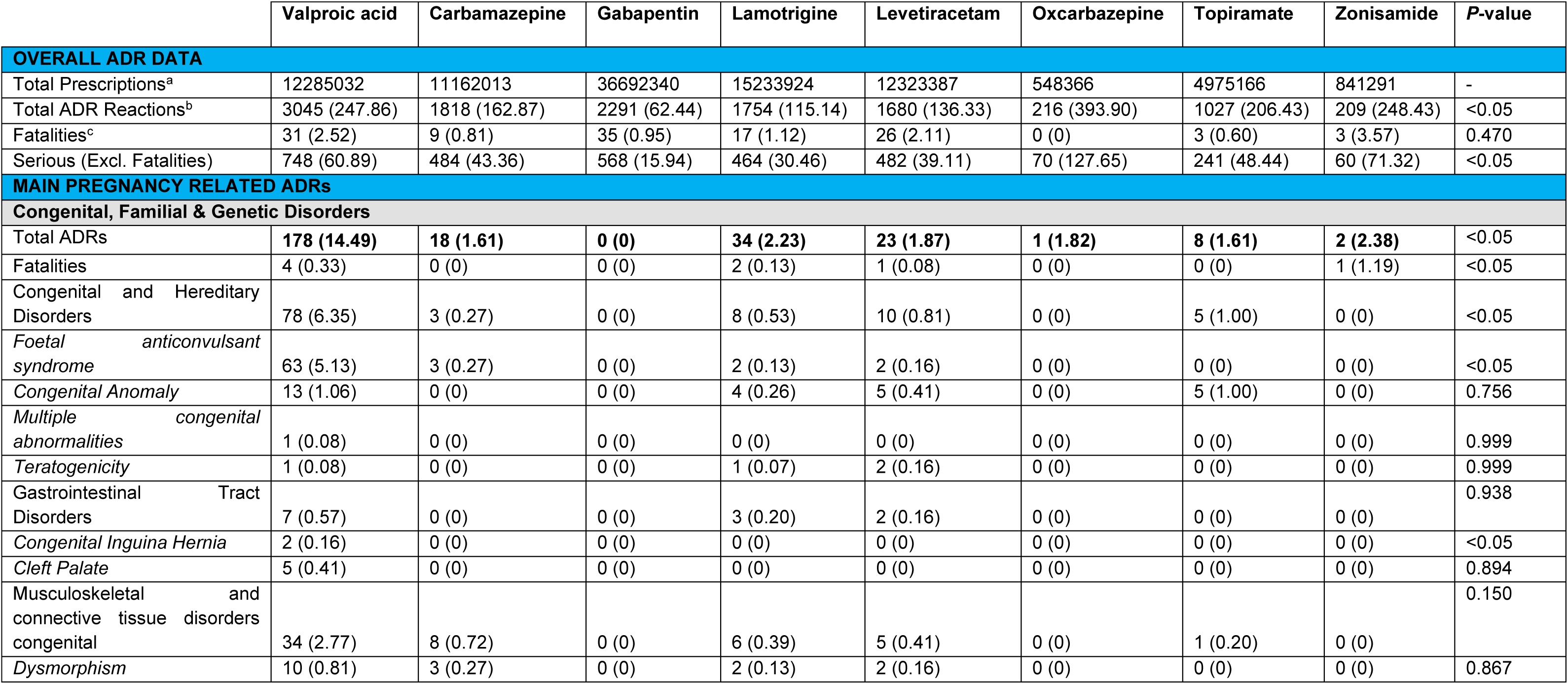
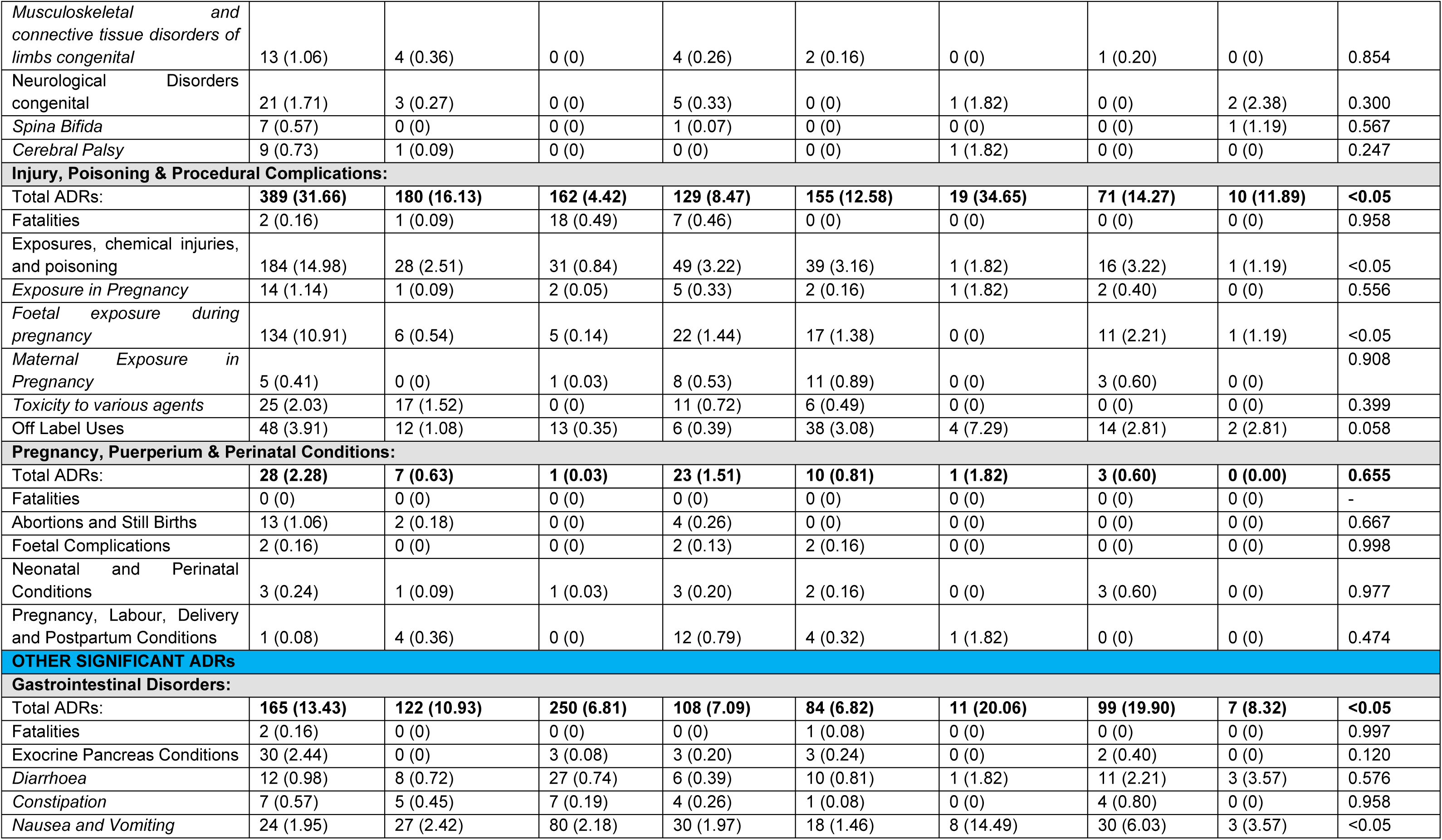
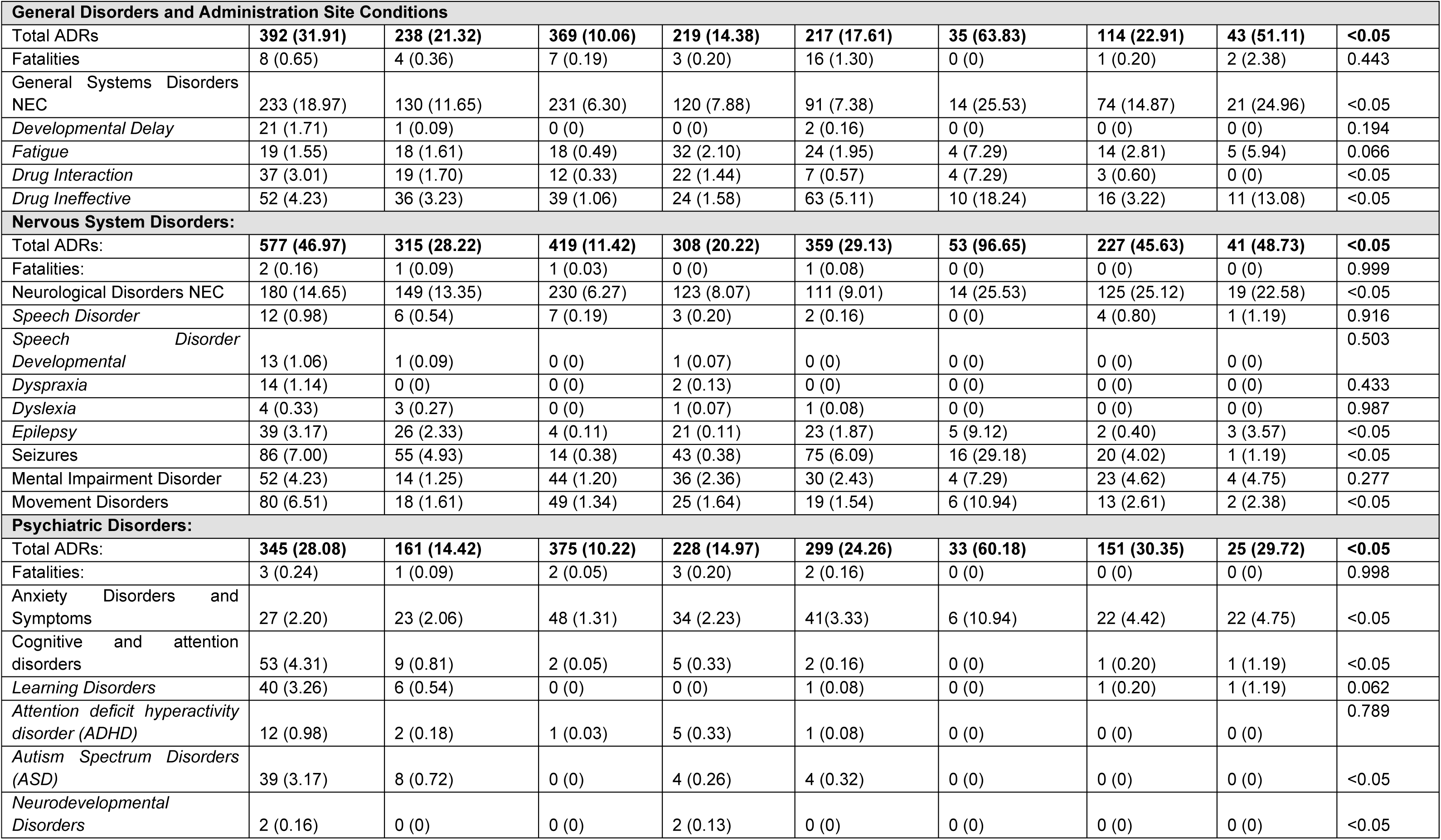
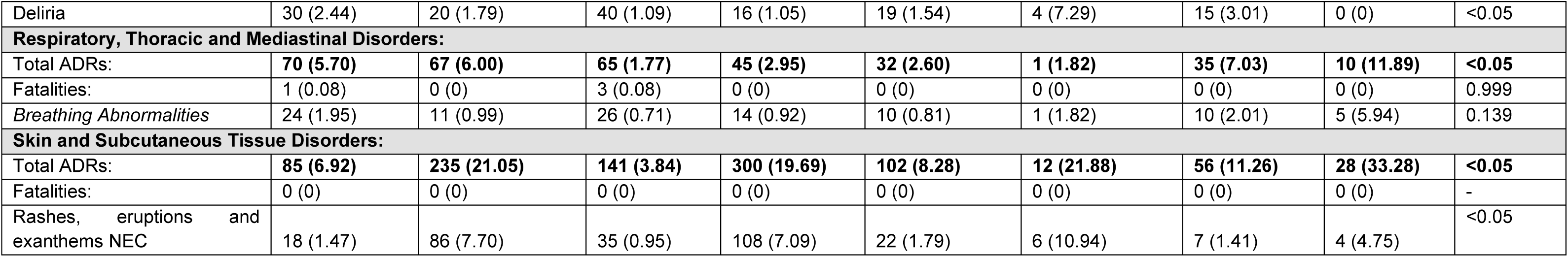
Summary of the reported suspected adverse drug reactions within the UK for all 8 studied AEDs. The number in parentheses represents the reported ADRs per 1,000,000 prescriptions as a standardised value for comparison. *P*-values were calculated through chi-squared analysis on the standardised data values. Key: Further details can be found in ^a^ **Chart S1**; ^b^ **Chart S2**; ^c^ **Chart S3** and **Figures S2-S9**.

### Congenital, Familial and Genetic Disorders

Valproic acid had the most suspected total ADRs in the congenital, familial, and genetic disorder category (14.49 per 1,000,000 *R_x_*). Valproic acid had the highest standardised rate of foetal anticonvulsant syndrome (5.13 per 1,000,000 *R_x_*). Zonisamide reported 1.19 fatalities per 1,000,000 *R_x_*, valproic acid (0.33), lamotrigine (0.13) and levetiracetam (0.08), respectively. Congenital, familial, and genetic disorder ADRs were suspected of association with valproic acid (14.49 per 1,000,000 *R_x_,* χ^2^ test, *P* < .05, *d* = 6.432, 95% CI [4.955, 8.348]). For congenital and hereditary disorders, valproic acid was unique (*d* = 9.069, 95% CI [5.807, 14.163]) (**Figure 1a**).

**Figure 1.**
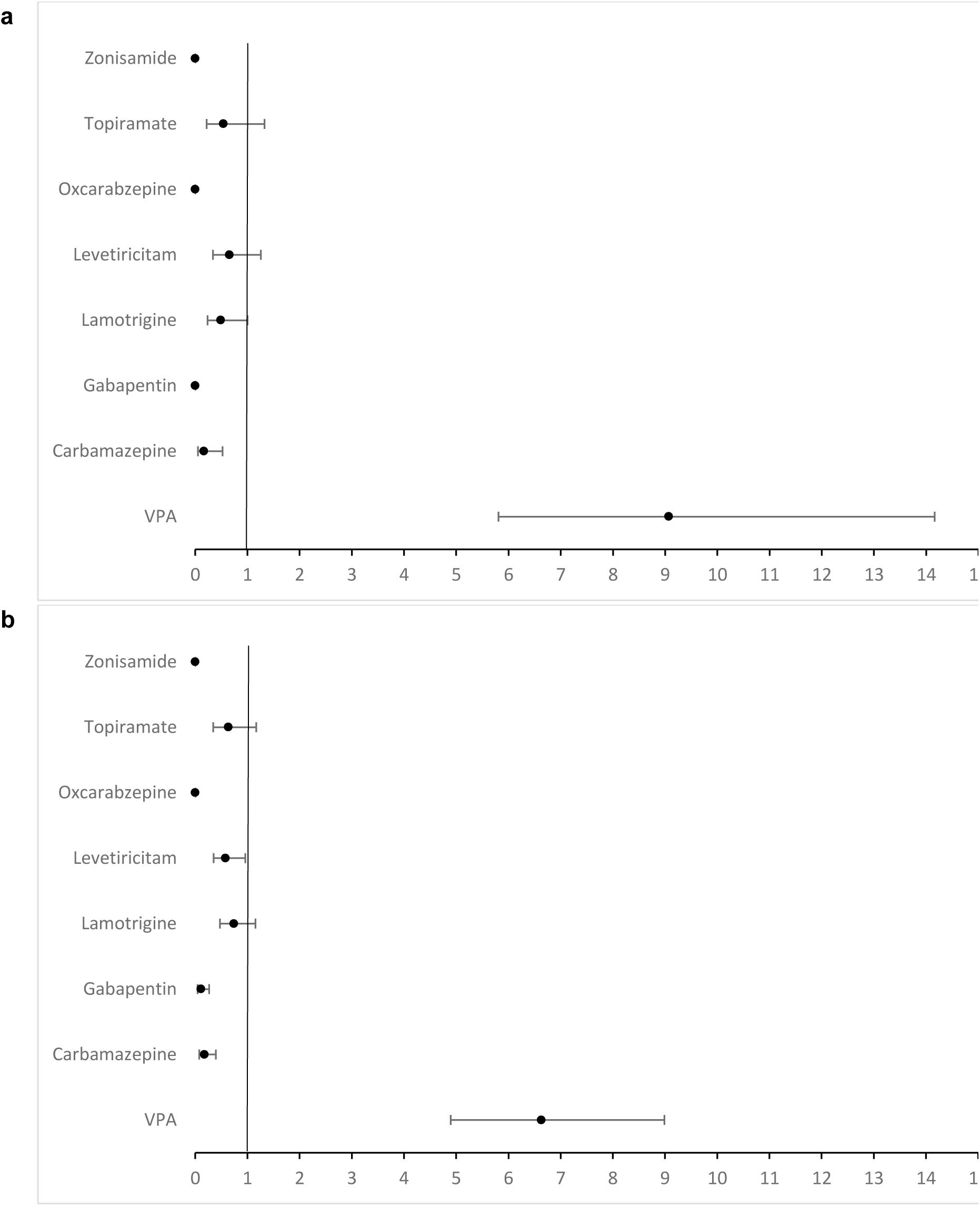
(**a**). Odds ratio and confidence interval for congenital and hereditary disorders for all AEDs studied; and **(b)** odds ratio and confidence interval for foetal exposure during pregnancy for all AEDs studied.

### Paternal route to ADRs

**Table S5** shows 5 reports have been made since 1972 for children born with congenital abnormalities due to suspected exposure to VPA via the father. However, due to transgenerational aspect of this data, it should be considered that there could be a significant degree of underreporting.

### Injury, Poisoning and Procedural Complications

Oxcarbazepine had the most suspected injury, poisoning, and procedural complication ADRs (34.65 per 1,000,000 *R_x_*), followed by valproic acid (31.66). Suspected foetal exposure during pregnancy was statistically significant across the AEDs (χ^2^, *P* < .05), with valproic acid reporting 10.91 per 1,000,000 *R_x_*, topiramate (2.21), lamotrigine (1.44), levetiracetam (1.38), zonisamide (1.19), carbamazepine (0.54), gabapentin (0.14) and oxcarbazepine (0.0). Gabapentin reported the most fatalities (0.49 per 1,000,000 *R_x_*). Injury, poisoning & procedural complications ADRs were suspected of association with valproic acid (31.66 per 1,000,000 *R_x_,* χ^2^ test, *P* < .05, *d* = 1.668, 95% CI [1.464, 1.901]), and carbamazepine (16.13 per 1,000,000 *R_x_,* χ^2^ test, *P* < .05, *d* = 1.091, 95% CI [0.923, 1.291]). For foetal exposure during pregnancy, valproic acid was unique (*d* = 6.632, 95% CI [4.894, 8.988] (**Figure 1b**).

### Gastrointestinal disorders

Oxcarbazepine had the most suspected gastrointestinal (GI) ADRs (20.06 per 1,000,000 *R_x_*) with nausea and vomiting the highest within this category (14.49 per 1,000,000 *R_x_*). Suspected fatalities due to drug-GI effects were reported for valproic acid (0.16 per 1,000,000 *R_x_*) and levetiracetam (0.08 per 1,000,000 *R_x_*). Gastrointestinal disorders ADRs were suspected of association with gabapentin (6.81 per 1,000,000 *R_x_,* χ^2^ test, *P* < .05, *d* = 1.881, 95% CI [1.611, 2.197]) and topiramate (19.90 per 1,000,000 *R_x_,* χ^2^ test, *P* < .05, *d* = 1.466, 95% CI [1.176, 1.827]).

### General disorders and administrative site conditions

Oxcarbazepine reported the most suspected general disorders and administrative site conditions (63.83 per 1,000,000 *R_x_*). Developmental delays were reported in valproic acid (1.71 per 1,000,000 *R_x_*), levetiracetam (0.16) and carbamazepine (0.09). Zonisamide reported the most fatalities (2.38 per 1,000,000 *R_x_*). General disorders and administration site conditions ADRs were suspected of association with zonisamide (51.11 per 1,000,000 *R_x_,* χ^2^ test, *P* < .05, *d* = 1.676, 95% CI [1.193, 2.353]), oxcarbazepine (63.83 per 1,000,000 *R_x_,* χ^2^ test, *P* < .05, *d* = 1.243, 95% CI [0.862, 1.792]), and gabapentin (10.06 per 1,000,000 *R_x_,* χ^2^ test, *P* < .05, *d* = 1.296, 95% CI [1.142, 1.470]).

### Nervous system disorders

Oxcarbazepine reported the most ADRs for epilepsy (9.12 per 1,000,000 *R_x_*) and seizures (29.18 per 1,000,000 *R_x_*). Fatalities were reported in four AEDs (valproic acid, carbamazepine, gabapentin, and levetiracetam) with valproic acid having the highest rate (0.16 per 1,000,000 *R_x_*). Nervous system disorders ADRs were suspected of association with oxcarbazepine (96.65 per 1,000,000 *R_x_,* χ^2^ test, *P* < .05, *d* = 1.387, 95% CI [1.014, 1.897]), topiramate (45.63 per 1,000,000 *R_x_,* χ^2^ test, *P* < .05, *d* = 1.224, 95% CI [1.049, 1.430]), levetiracetam (29.13 per 1,000,000 *R_x_,* χ^2^ test, *P* < .05, *d* = 1.180, 95% CI [1.039, 1.339]), and zonisamide (48.73 per 1,000,000 *R_x_,* χ^2^ test, *P* < .05, *d* = 1.035, 95% CI [0.733, 1.460]).

### Psychiatric Disorders

Oxcarbazepine had the highest suspected rate of psychiatric ADRs (60.18 per 1,000,000 *R_x_*). The size effect of these suspected psychiatric disorder ADRs were as follows: levetiracetam (24.26 per 1,000,000 *R_x_,* χ^2^ test, *P* < .05, *d* = 1.485, 95% CI [1.294, 1.705]), gabapentin (10.22 per 1,000,000 *R_x_,* χ^2^ test, *P* < .05, *d* = 1.341, 95% CI [1.182, 1.520]), oxcarbazepine (60.18 per 1,000,000 *R_x_,* χ^2^ test, *P* < .05, *d* = 1.166, 95% CI [0.802, 1.695]), and topiramate (30.35 per 1,000,000 *R_x_,* χ^2^ test, *P* < .05, *d* = 1.123, 95% CI [0.936, 1.346]). Anxiety disorders and symptoms were the most reported in oxcarbazepine, zonisamide, topiramate, levetiracetam, lamotrigine, valproic acid, carbamazepine and gabapentin in a descending order. Valproic acid had the most reports for learning disorders (3.26 per 1,000,000 *R_x_*), attention-deficit hyperactivity disorder [ADHD] (0.98 per 1,000,000 *R_x_*) and autism spectrum disorders [ASD] (3.17 per 1,000,000 *R_x_*).

### Other ADRs

#### Pregnancy, Puerperium & Perinatal Conditions ADRs

Pregnancy, Puerperium & Perinatal (PPP) conditions ADRs were suspected of association with lamotrigine (1.51 per 1,000,000 *R_x_,* χ^2^ test, *P* < .05, *d* = 2.720, 95% CI [1.656, 4.469]), and valproic acid (2.28 per 1,000,000 *R_x_,* χ^2^ test, *P* < .05, *d* = 1.846, 95% CI [1.150, 2.964]). PPP ADRs include the following MedDRA terms: abortions and still births, foetal complications, neonatal and perinatal conditions, and pregnancy, labour, delivery and postpartum conditions in this study.

#### Respiratory, Thoracic and Mediastinal Disorders ADRs

Respiratory, thoracic, and mediastinal disorders were most reported in zonisamide (11.89 per 1,000,000 *R_x_*). Gabapentin and valproic acid reported fatalities (both 0.08 per 1,000,000 *Rx*). Respiratory, Thoracic and Mediastinal Disorders ADRs were suspected of association with zonisamide (11.89 per 1,000,000 *R_x_,* χ^2^ test, *P* < .05, *d* = 1.837, 95% CI [0.964, 3.501]), carbamazepine (6.00 per 1,000,000 *R_x_,* χ^2^ test, *P* < .05, *d* = 1.478, 95% CI [1.124, 1.943]), topiramate (7.03 per 1,000,000 *R_x_,* χ^2^ test, *P* < .05, *d* = 1.305, 95% CI [0.913, 1.864]), and gabapentin (1.77 per 1,000,000 *R_x_,* χ^2^ test, *P* < .05, *d* = 1.066, 95% CI [0.809, 1.404]).

#### Skin and Subcutaneous Tissue Disorders ADRs

Zonisamide had the most skin and subcutaneous tissue disorder ADRs (33.28 per 1,000,000 *R_x_*), with oxcarbazepine reporting the most rashes (10.94 per 1,000,000 *R_x_*). No fatalities were reported. Skin and subcutaneous tissue disorders ADRs were suspected of association with lamotrigine (19.69 per 1,000,000 *R_x_,* χ^2^ test, *P* < .05, *d* = 3.014, 95% CI [2.601, 3.492]), carbamazepine (21.05 per 1,000,000 *R_x_,* χ^2^ test, *P* < .05, *d* = 1.948, 95% CI [1.665, 2.277]), and zonisamide (33.28 per 1,000,000 *R_x_,* χ^2^ test, *P* < .05, *d* = 1.811, 95% CI [1.210, 2.712]).

## Discussion

The 8 AEDs have differing modes of action. Valproic acid increases GABAergic (g-aminobutyric acid) and glutamatergic neurotransmission while inhibiting neuronal sodium channels and altering cellular signalling and ERK pathways. Valproic acid is a potent first-generation antiepileptic but is known to be teratogenic and can cause liver and pancreas damage.[34–36] Carbamazepine, topiramate, lamotrigine, oxcarbazepine and zonisamide increase GABA transmission and block voltage-gated sodium channels to inhibit neuronal firing and suppress the release of excitatory amino acid glutamate. Levetiracetam causes modulation of synaptic neurotransmission release by binding to synaptic vesicle protein SV2A which can inhibit neurotransmitter release.[37] Gabapentin inhibits α-2-δ subunit of voltage-gated calcium channels resulting in glutamate release. The polypharmacology of these AEDs may explain the ADRs observed. Secondly, it is possible that either the beneficial effects or the adverse effects, perhaps both, are produced by different combinations of mechanisms, some of which are labelled “off-targets”.

AEDs control seizure activity through disparate mechanisms which may elicit different ADR profiles.[38] oxcarbazepine had the most suspected ADRs (393.90 per 1,000,000 *R_x_*) compared to gabapentin with the least (62.44). Oxcarbazepine and zonisamide are the least prescribed and newest AEDs.

It was expected that the black triangle drug, valproic acid would have the most off-target interactions based on LLE <5. Clinically achievable targeting of HDACs by valproic acid is associated with teratogenicity (HDAC 1 IC_50_ = 54.4 µM, HDAC2 IC_50_ = 82.4 µM, HDAC3 IC_50_ = 148 µM, HDAC8 IC_50_ = 144 µM, valproic acid C_max_ = 184.3 µM). valproic acid was the only AED with an odds value >>1 for foetal exposure ((*d* = 6.632, 95% CI [4.894, 8.988]) and congenital and hereditary disorders (valproic acid (*d* = 9.069, 95% CI [5.807, 14.163]).

### All ADRs

Oxcarbazepine had the most standardised reported suspected ADRs for psychiatric (60.18 per 1,000,000 *R_x_*) and nervous system disorders (69.65). Suspected skin reactions were reported most with zonisamide, potentially linked to its ability to induce lupus through increasing anti-nucleic antibodies.[39] Valproic acid is known to cause pancreatitis and displayed the highest relative rate of exocrine pancreatic conditions (2.44 per 1,000,000 *R_x_*).[40] Topiramate had similar standardised suspected ADRs to other AEDs, with most suspected reactions in psychiatric disorders (30.35 per 1,000,000 *R_x_*) and nervous system (45.63). Paraesthesia, caused by inhibition of carbonic anhydrase enzymes (carbonic anhydrase I IC_50_ = 250 nM and carbonic anhydrase II IC_50_ = 868.3 nM *vs.* C_max_ = 4420 nM [topiramate]) alters nerve cell communication and/or its effects on peripheral nerve cells.[41]

Carbamazepine had suspected nervous system disorders (21.32 per 1,000,000 *R_x_*), including neurodevelopmental disorders which may be caused by histone deacetylase targeting (HDAC3 and 7).[42] Skin disorders (21.05 per 1,000,000 *R_x_*) may be explained by carbamazepine rapidly activating T-cells through direct interaction with major histocompatibility (MHC) proteins and specific T-cell receptors causing an immune response.[43]

Levetiracetam was suspected of nervous system (*d* = 1.180 [1.039-1.339]) and psychiatric disorder ADRs (*d* = 1.485 [1.294-1.705]) and displayed one of the higher reported seizure rates (6.09 per 1,000,000 *R_x_*), suggesting a lower level of epilepsy control.[44]

Lamotrigine’s suspected ADRs were in pregnancy or skin conditions, with lamotrigine-induced rash a well-reported side effect, with hypersensitivity from valproic acid interfering with glucuronide metabolism and increasing lamotrigine blood levels.[45] *h*ERG was targeted by lamotrigine and carbamazepine but not at levels associated with arrythmia.

Gabapentin had the lowest standardised ADRs. Gastrointestinal, psychiatric, and GI disorders were the predominant ADRs and its ability to target α-2-δ voltage-gated calcium channels inhibit neurotransmitter release but causes glutamate-induced glutamate release from astrocytes generating neurotoxicity and psychiatric symptoms.[46]

### Congenital, Familial and Genetic Disorder ADRs and Neurodevelopmental ADRs

All AEDs except gabapentin reported congenital, familial, and genetic disorder ADRs with statistical significance for valproic acid (**Table S3**). Valproic acid was the only AED to display a strong association between exposure and this ADR, with *d* = 6.432 [4.955-8.348] (**Figure 1a**).

Foetal exposure ADR is most associated with valproic acid (**Table 2**). Valproic acid crosses the placental barrier with increasing concentrations in cord blood than maternal serum, higher doses increase foetal accumulation, increasing the pharmacological effect.[47] Studies have linked teratogenicity to direct inhibition of placental folate uptake through blocking folate receptors, specifically FOLR1. FOLR1 affects embryonic folate metabolism through inhibiting glutamate formyl transferase while increasing plasma homocysteine levels and reducing serum folate, causing placental folate deficiency which limits synthesis of purines and thymine required for DNA formation, resulting in congenital malformations.[12,48–50]

Valproic acid is suspected of causing neurodevelopmental disorder ADRs, including learning disorders (3.26 per 1,000,000 *R_x_*), autism (3.17), developmental delay (1.71), developmental speech disorders (1.06), and ADHD (0.98). This may be linked to valproic acid’s ability to inhibit HDAC2 (IC_50_ = 82.4 µM, C_max_ = 184.3 µM).[42] HDAC2 inhibition affects cellular differentiation, causes apoptosis leading to congenital, and developmental disorders through histone hyperacetylation, affecting normal gene transcription.[51] *In-utero* exposure causes atrial septal defect (ASD) through transient histone hyperacetylation in the embryonic brain, increasing apoptotic cells in the neocortex and decreasing cell proliferation while altering *m*RNA levels, delaying neuronal maturation.[42]

Carbamazepine is considered teratogenic.[52] Off-target interactions with HDAC3 and 7 (both IC_50_ = 2 µM, C_max_ = 11-14.8 µM) may contribute to suspected congenital, familial and genetic disorders ADRs (1.61 per 1,000,000 *R_x_*).[53] It disturbs cardiac rhythm secondary to its propensity to inhibit specific ion current (Ikr) and subsequent hypoxic damage as Ikr is essential for embryonic cardiac repolarisation and rhythm regulation and impairs folate absorption.[54] Topiramate has an identical suspected congenital, familial and genetic disorder ADRs profile to carbamazepine and increased congenital and neurodevelopmental disorders, risk of low birth weight and foetal growth restrictions. This is due to HDAC inhibition causing hyperacetylation in human cells at lower potency than valproic acid. Topiramate inhibits carbonic anhydrase (I and II, IC_50_ = 0.25 and 0.87 µM, respectively *vs.* C_max_ = 4.4 µM) which reduces embryonic intracellular *p*H but is required to control cellular development.[54]

Zonisamide has limited studies into pregnancy ADRs (2.38 per 1,000,000 *R_x_*), however carbonic anhydrase inhibition (II and V, 35.2 and 20.6 nM, respectively *vs.* C_max_ = 9.4 µM) may cause teratogenicity. It causes low birth weight for gestational age in 21% of exposure and animal models support its teratogenic potential.[55]

Lamotrigine (2.23 per 1,000,000 *R_x_*) and levetiracetam (1.87 per 1,000,000 *R_x_*) had higher standardised congenital, familial, and genetic disorder ADRs than carbamazepine and topiramate (both 1.61 per 1,000,000 *R_x_*) despite being considered safer AEDs in pregnancy.[8] Lamotrigine dual therapy (with vigabatrin) has been reported to increase neurodevelopmental risk and its effect on dihydrofolate reductase (DHFR) may be teratogenic.[54,56]

Levetiracetam’s major metabolite, 2-pyrrolidinone-*n*-butyric acid (PBA), is an unspecified HDAC class inhibitor which can be teratogenic, however studies support a margin of reproductive safety.[57] Oxcarbazepine shows congenital malformations (1.82 per 1,000,000 *R_x_*) but insufficient maternal exposure prevents conclusions being drawn and studies are limited into teratogenic mechanisms.[58]

Overall, suspected teratogenic effects were shown to occur with AEDs but are relatively uncommon, except for valproic acid, and can be linked to their pharmacological activity. Targeting of carbonic anhydrase or histone deacetylase can result in specific receptor-mediated teratogenesis and are the most likely causes of these ADRs occurring alongside some AED’s role in folate antagonism causing foetal folate deficiency.[54] Valproic acid had the most congenital and neurodevelopmental adverse effects, potentially linked to it inhibiting the most histone deacetylase isozymes and its interference in folate metabolism.[54] Other AEDs which inhibit carbonic anhydrase included carbamazepine, topiramate and levetiracetam’s active metabolite PBA – all reported congenital and neurodevelopmental adverse reactions. The teratogenic effects of lamotrigine and oxcarbazepine are unknown, however both drugs have reported congenital and neurodevelopmental ADRs. Lamotrigine, valproic acid and carbamazepine all undergo folate antagonism which is known to be a further teratogenic mechanism by inhibiting the folate methylation cycle, with carbamazepine able to impair folate absorption, lamotrigine inhibiting DHFR, and valproic acid being a folate anti-metabolite.[54]

### Exposure via the father

The route of exposure of a foetus to sodium valproate is highly topical at the time of this work. It is known that valproic acid can affect a foetus *in utero* by exposure via the mother during pregnancy, but it is unknown whether exposure via the father could be a potential cause of NTDs and neurological conditions. A study found that hyperacetylation of testicular histone can be linked to trans-generational genetic changes.[59] Impaired DNA methylation was linked to impaired promoter CpGs of genes which are related to brain function. This could suggest that neurological changes can be inflicted on the foetus due to modifications of the gamete. The pattern of mutations in the sperm found that many mutations were found near acetylated histone regions, suggesting valproic acid influences methylation, although not necessarily directly. The implications of these findings were changes to national guidance in the UK advising against valproic acid prescriptions in any person under 55, extending this guidance to males.[60] The FOI request (**Table S5**) identified 5 suspected reports of NTDs or neurological abnormalities due to VPA exposure via the father since 1972. However, due to the nature of the changes, the trans-generational ADRs could be at risk of being under-reported. Neurological and behavioural changes such as ASD may not present until much later in a child’s development and a link to exposure to VPA as a sperm cell could plausibly be missed.

## Limitations

Under-reporting of suspected ADRs prevents accurate quantification of the risks associated with AEDs.[61] Suspected ADR reporting requires no causality to be determined and co-morbidities and polypharmacy of patients is not publicly accessible information. Neurodevelopmental disorders have overlapping symptoms and can take years to be diagnosed, with adolescent and adult diagnosis common, meaning links between AED use in pregnancy may be missed.[62] Different ADRs are likely to be under-reported at widely different rates, especially teratogenic events, which are much more likely to be reported than other events, thus data reported by the UK’s National Congenital Anomaly and Rare Disease Registration Service (NCARDRS) is used for comparison.[63] Some congenital defects are reported on behalf of the child and some on behalf of the mother who gave birth to the affected child. It was not possible to filter by route of exposure because some congenital defects are reported on behalf of the child (which would be trans-placental) and some on behalf of the mother (which would be oral) who gave birth to the affected child, so only searches for congenital defects which occurred *in-utero* was possible. A consistent, nationally implemented method of reporting ADRs is required, to have a better understanding of the clinical application of these findings, and to allow for the most effective and appropriate AED to be prescribed.

Valproic acid is more widely prescribed in men and over 55s than it is in women of childbearing age due to the restrictions in prescribing valproic acid to women, meaning the number of congenital defects as a percentage of female patients (of childbearing age) is again likely to be higher. It could be hypothesised that this would also be true when comparing valproic acid to lamotrigine and carbamazepine, two drugs without the same restrictions in prescribing to women. Lamotrigine and carbamazepine also do not have a PPP, making the data less comparable.

Due to transgenerational aspect of this data, there could be a significant degree of underreporting, especially for conditions such as ASD where symptoms do not appear until later in life. The ambiguity of valproic acid exposure via the father could further contribute to this.

OpenPrescribing gives 5-year prescribing data for England (83.9% of the UK population in mid-2021), whereas ADR data is for the UK, causing standardisation to appear higher than the true value by approximately 19.2%. This can only be an approximation as a complex set of factors influencing ADRs, including social deprivation and drug and alcohol use, vary between regions. Prescribing guidance was implemented in 2018 (the PPP) for the black triangle drug valproic acid therefore some ADR reports will include valproic acid exposure *in-utero* prior to this.

Of the eight AEDs studied, only valproate is a black-triangle drug in the UK. That suggests that suspected ADRs to valproate would have been more likely to have been reported than suspected ADRs to the other drugs, thus potentially skewing the data.

Standardisation per 1,000,000 Rx improves validity due to the need for n ≥ 5 reports for accurate statistical analysis. Not all organ systems met ≥ 5 reported reactions required for odds ratio calculations. Incomplete pharmacological profiles would also limit potential mechanistic correlations.

## Conclusion

All AEDs examined in this study have unique suspected ADR profiles. ADRs per 1,000,000 *R_x_* identified across all AEDs are statistically significant (χ^2^ test, *P* < .05).

Pregnancy, puerperium & perinatal conditions ADRs suspected of association with lamotrigine (1.51 per 1,000,000 *R_x_,* χ^2^ test, *P* < .05, *d* = 2.720, 95% CI [1.656, 4.469]) had a larger size effect than valproic acid (2.28 per 1,000,000 *R_x_,* χ^2^ test, *P* < .05, *d* = 1.846, 95% CI [1.150, 2.964]).

All AEDs studied except for gabapentin had teratogenic effects. With at least one reported congenital, familial, and genetic disorder ADR, primarily foetal anticonvulsant syndrome or congenital anomaly. The size effect associated with valproic acid for congenital and hereditary disorders, (*d* = 9.069, 95% CI [5.807, 14.163]) and foetal exposure during pregnancy, the size effect associated with valproic acid (*d* = 6.632, 95% CI [4.894, 8.988]) were pronounced. Valproic acid had polypharmacology including the unique and clinically achievable targeting of histone deacetylase (HDAC 1 IC_50_ = 54.4 µM, HDAC2 IC_50_ = 82.4 µM, HDAC3 IC_50_ = 148 µM, HDAC8 IC_50_ = 144 µM, C_max_ = 184.3 µM) associated with teratogenicity.

Consideration of the paternal exposure route to valproic acid birth defects revealed five suspected reports. However, these are likely underreported clinically due to its trans-generational aspect and the delay in the appearance of behavioural/neurological symptoms the link may not be made to valproic acid exposure as a gamete.

## Acknowledgements

The authors thank the MHRA Yellow Card, ChEMBL, OpenPrescribing for access to the underlying data for this study.

## Data availability statements

All supporting data is freely available via the supporting information file(s).

## Author contribution statement

AMJ and VS designed the project, supervised, and drafted/revised the manuscript, BP and IE carried out the investigation and statistical analyses and drafted the manuscript.

## Ethics approval statement

The study used fully anonymised publicly available data and ethics approval was not required.

