## Supplementary Information for "United Kingdom National Register Study of Anti-Epileptic Medications: Suspected Foetal Congenital And Pregnancy-Associated Side Effects"

### CONTENTS


| | |
|---|---|
| **Figure S1.** Chemical structures of the AEDs studied. | **Page 2** |
| **Table S1:** Inclusion/Exclusion criteria for each AED. | **Page 3** |
| **Table S2:** Excluded AEDs. | **Page 4** |
| **Equation S1.** ADR rate standardization to prescribing levels | **Page 5** |
| **Equation S2.** OR/CI calculations. | **Page 6** |
| **Table S3:** Statistical summary of the eight AEDs ADRs compared individually to each other (Chi-squared test). | **Pages 7-9** |
| **Table S4:** The number and type of ADR reports for routes of exposure related to the father (data is from a FOI response from the MHRA regarding foetal exposure to VPA via the father 27/10/2023) | **Page 10** |
| **Chart S1:** Number of Prescriptions dispensed per AED between August 2017 – July 2022. | **Page 11** |
| **Chart S2:** Number of standardised reported AED ADRs per 1 million prescriptions between January 2018-August 2022. | **Page 12** |
| **Chart S3:** Total number of standardised reported suspected fatalities per 1 million prescriptions between January 2018-August 2022. | **Page 13** |
| **Figure S2.** Odds ratio and confidence interval for VPA. | **Page 14** |
| **Figure S3.** Odds ratio and confidence interval for carbamazepine. | **Page 15** |
| **Figure S4.** Odds ratio and confidence interval for gabapentin. | **Page 16** |
| **Figure S5.** Odds ratio and confidence interval for lamotrigine. | **Page 17** |
| **Figure S6.** Odds ratio and confidence interval for levetiracetam. | **Page 18** |
| **Figure S7.** Odds ratio and confidence interval for oxcarbazepine. | **Page 19** |
| **Figure S8.** Odds ratio and confidence interval for topiramate. | **Page 20** |
| **Figure S9.** Odds ratio and confidence interval for zonisamide. | **Page 21** |
| **Table S5:** Physiochemical and pharmacokinetic properties of the eight AEDs studied. | **Pages 22-23** |

**Figure S1.** Chemical structures of the AEDs studied.

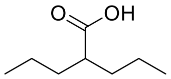

valproic acid (VPA)

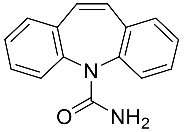

carbamazepine

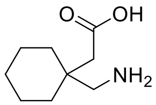

gabapentin

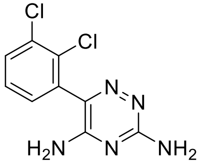

lamotrigine

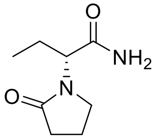

levetiracetam

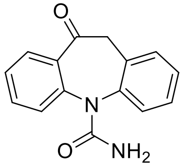

oxcarbazepine

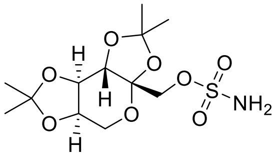

topiramate

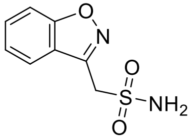

zonisamide

**Table S1:** Inclusion/Exclusion criteria for each AED.

| INCLUSION | EXCLUSION |
|---|---|
| • Drug's 5-year prescribing data availability.<br>• Human single protein polypharmacology data (n ≥ 2 targets) for the drug was available.<br>• ADR Data available for 5-year window.<br>• Drug is a UK MHRA licensed anti-epileptic (pre-2017).<br>• Drug's main indication was for epilepsy. | • Drug has <500,000 Prescriptions during the 5-year period.<br>• Human single protein polypharmacology data (n ≤ 1 targets) for the drug.<br>• Incomplete ADR Data availability for the 5-year period.<br>• Unlicensed for the indication of epilepsy in the UK.<br>• Uncommonly used for epilepsy, with a different main indication, e.g., pain.<br>• Previous recall of any batches.<br>• Full details in supporting information (Table **S1**) |

**Table S2:** Excluded AEDs

| AED Name: | Total Prescriptions: | Total Reactions: | Fatalities: | Main reason for Exclusion: |
|---|---|---|---|---|
| Brivaracetam | 181666 | 406 | 8 | <500,000 Prescriptions. |
| Cannabidiol | 16 | 699 | 1 | <500,000 Prescriptions. |
| Cenobamate | 236 | 20 | 0 | <500,000 Prescriptions and first ADR reported 2021. |
| Eslicarbazepine Acetate | 115803 | 58 | 0 | <500,000 Prescriptions. |
| Ethosuximide | 194700 | 58 | 0 | <500,000 Prescriptions. |
| Fenfluramine | 0 | 14 | 0 | <500,000 Prescriptions. |
| Fosphenytoin Sodium | 3 | - | - | <500,000 Prescriptions and no ADR data since 2012. |
| Lacosamide | 870436 | 385 | 3 | Quality defect leading to recall in 2011 and thereafter.[103, 104] |
| Peramapanel | 253129 | 260 | 2 | <500,000 Prescriptions. |
| Phenytoin | 102498 | 822 | 14 | <500,000 Prescriptions. |
| Pregablin | 38001388 | 3874 | 144 | Main indication not Epilepsy. |
| Rufinamide | 61955 | 31 | 1 | <500,000 Prescriptions. |
| Stiripentol | 13703 | 18 | 1 | <500,000 Prescriptions. |
| Tiagabine | 8397 | 3 | 0 | <500,000 Prescriptions and no ADR reports since 2019. |
| Vigabatrin | 66184 | 25 | 0 | <500,000 Prescriptions. |

**Equation S1.** ADR rate standardization to prescribing levels

$$Standardised\ ADR\ Rate = \frac{Total\ Reported\ Reactions}{Total\ Prescriptions}\ x\ 1{,}000{,}000$$

**Equation S2.** OR/CI calculations.

| | Target Drug | All other AED's |
|---|---|---|
| Target ADR | a | b |
| All other ADRs | c | d |
| | a/b | c/d |

$$Odds\ Ratio = \frac{(a/b)}{(c/d)}$$

$$95\%\ Confidence\ Interval = e^{ln(ROR)\pm 1.96\sqrt{(\frac{1}{a}+\frac{1}{b}+\frac{1}{c}+\frac{1}{d})}}$$

**Table S3:** Statistical Summary of the eight AEDs ADRs compared individually to each other (Chi-Squared Test):

| | Studied Data | Valproic Acid V | Valproic Acid V | Valproic Acid V | Valproic Acid V | Valproic Acid V | Valproic Acid V Topiramate | Valproic Acid V | Carbamazepine V | Carbamazepine V | Carbamazepine | Carbamazepine V | Carbamazepine V | Carbamazepine V | Gabapentin V | Gabapentin V | Gabapentin V | Gabapentin V Topiramate | Gabapentin V | Lamotrigine V | Lamotrigine V | Lamotrigine V Topiramate | Lamotrigine V | Levetiracetam V | Levetiracetam V | Levetiracetam V | Oxcarbazepine V | Oxcarbazepine V | Topiramate V Zonisamide |
|---|---|---|---|---|---|---|---|---|---|---|---|---|---|---|---|---|---|---|---|---|---|---|---|---|---|---|---|---|---|
| Total ADR Reactions | <0.05 | <0.05 | <0.05 | <0.05 | <0.05 | <0.05 | 0.052 | 0.980 | <0.05 | <0.05 | 0.125 | <0.05 | <0.05 | <0.05 | <0.05 | <0.05 | <0.05 | <0.05 | <0.05 | 0.181 | <0.05 | <0.05 | <0.05 | <0.05 | <0.05 | <0.05 | <0.05 | <0.05 | <0.05 |
| Fatalities | 0.470 | 0.347 | 0.400 | 0.461 | 0.848 | 0.112 | 0.277 | 0.673 | 0.911 | 0.823 | 0.445 | 0.369 | 0.864 | 0.187 | 0.910 | 0.509 | 0.329 | 0.779 | 0.219 | 0.580 | 0.291 | 0.696 | 0.258 | 0.146 | 0.360 | 0.541 | 0.437 | 0.059 | 0.147 |
| Serious (Excl. Fatalities) | <0.05 | 0.086 | <0.05 | <0.05 | <0.05 | <0.05 | 0.234 | 0.364 | <0.05 | 0.133 | 0.640 | <0.05 | 0.596 | <0.05 | <0.05 | <0.05 | <0.05 | <0.05 | <0.05 | 0.299 | <0.05 | <0.05 | <0.05 | <0.05 | 0.319 | <0.05 | <0.05 | <0.05 | <0.05 |
| **MAIN PREGNANCY RELATED ADRs** | | | | | | | | | | | | | | | | | | | | | | | | | | | | | |
| **Congenital, Familial & Genetic Disorders** | | | | | | | | | | | | | | | | | | | | | | | | | | | | | |
| Total ADRs | <0.05 | <0.05 | <0.05 | <0.05 | <0.05 | <0.05 | <0.05 | <0.05 | 0.204 | 0.752 | 0.892 | 0.909 | 0.998 | 0.702 | 0.135 | 0.172 | 0.177 | 0.205 | 0.123 | 0.857 | 0.839 | 0.750 | 0.946 | 0.982 | 0.890 | 0.804 | 0.907 | 0.787 | 0.700 |
| Fatalities | <0.05 | 0.568 | 0.568 | 0.774 | 0.702 | 0.568 | 0.568 | 0.483 | - | 0.717 | 0.776 | - | - | 0.276 | 0.717 | 0.776 | - | - | 0.276 | 0.913 | 0.717 | 0.717 | 0.357 | 0.776 | 0.776 | 0.326 | - | 0.276 | 0.276 |
| Congenital and Hereditary Disorders | <0.05 | <0.05 | <0.05 | <0.05 | <0.05 | <0.05 | <0.05 | <0.05 | <0.05 | 0.774 | 0.602 | 0.604 | 0.514 | 0.604 | 0.469 | 0.368 | - | 0.316 | - | 0.804 | 0.469 | 0.698 | 0.469 | 0.368 | 0.886 | 0.368 | 0.316 | - | 0.316 |
| *Foetal anticonvulsant syndrome* | <0.05 | <0.05 | <0.05 | <0.05 | <0.05 | <0.05 | <0.05 | <0.05 | 0.604 | 0.828 | 0.871 | 0.604 | 0.604 | 0.604 | 0.717 | 0.687 | - | - | - | 0.954 | 0.717 | 0.717 | 0.717 | 0.687 | 0.687 | 0.687 | - | - | - |
| *Congenital Anomaly* | 0.756 | 0.304 | 0.304 | 0.489 | 0.590 | 0.304 | 0.970 | 0.304 | - | 0.608 | 0.524 | - | 0.316 | - | 0.608 | 0.524 | - | 0.316 | - | 0.861 | 0.608 | 0.510 | 0.608 | 0.524 | 0.614 | 0.524 | 0.316 | - | 0.316 |
| *Multiple congenital abnormalities* | 0.999 | 0.775 | 0.775 | 0.775 | 0.775 | 0.775 | 0.775 | 0.775 | - | - | - | - | - | - | - | - | - | - | - | - | - | - | - | - | - | - | - | - | - |
| *Teratogenicity* | 0.999 | 0.775 | 0.775 | 0.967 | 0.870 | 0.775 | 0.775 | 0.775 | - | 0.798 | 0.687 | - | - | - | 0.798 | 0.687 | - | - | - | 0.840 | 0.798 | 0.798 | 0.798 | 0.687 | 0.687 | 0.687 | - | - | - |
| Gastrointestinal Tract Disorders | 0.938 | 0.450 | 0.450 | 0.670 | 0.634 | 0.450 | 0.450 | 0.450 | - | 0.657 | 0.687 | - | - | - | 0.657 | 0.687 | - | - | - | 0.954 | 0.657 | 0.657 | 0.657 | 0.687 | 0.687 | 0.687 | - | - | - |
| *Congenital Inguina Hernia* | <0.05 | 0.687 | 0.687 | 0.687 | 0.687 | 0.687 | 0.687 | 0.687 | - | - | - | - | - | - | - | - | - | - | - | - | - | - | - | - | - | - | - | - | - |
| *Cleft Palate* | 0.894 | 0.523 | 0.523 | 0.523 | 0.523 | 0.523 | 0.523 | 0.523 | - | - | - | - | - | - | - | - | - | - | - | - | - | - | - | - | - | - | - | - | - |
| Musculoskeletal and connective tissue disorders congenital | 0.150 | 0.272 | 0.096 | 0.182 | 0.185 | 0.096 | 0.136 | 0.096 | 0.397 | 0.759 | 0.769 | 0.397 | 0.590 | 0.397 | 0.530 | 0.524 | - | 0.654 | - | 0.989 | 0.530 | 0.803 | 0.530 | 0.524 | 0.793 | 0.524 | 0.654 | - | 0.654 |
| *Dysmorphism* | 0.867 | 0.600 | 0.367 | 0.483 | 0.510 | 0.367 | 0.367 | 0.367 | 0.604 | 0.828 | 0.871 | 0.604 | 0.604 | 0.604 | 0.717 | 0.687 | - | - | - | 0.954 | 0.717 | 0.717 | 0.717 | 0.687 | 0.687 | 0.687 | - | - | - |
| *Musculoskeletal and connective tissue disorders of limbs congenital* | 0.854 | 0.557 | 0.304 | 0.489 | 0.417 | 0.304 | 0.445 | 0.304 | 0.549 | 0.903 | 0.786 | 0.549 | 0.833 | 0.549 | 0.608 | 0.687 | - | 0.654 | - | 0.878 | 0.608 | 0.928 | 0.608 | 0.687 | 0.949 | 0.687 | 0.654 | - | 0.654 |
| Neurological Disorders congenital | 0.300 | 0.306 | 0.191 | 0.333 | 0.191 | 0.952 | 0.191 | 0.741 | 0.604 | 0.939 | 0.604 | 0.282 | 0.604 | 0.195 | 0.567 | - | 0.177 | - | 0.123 | 0.567 | 0.308 | 0.567 | 0.213 | 0.177 | - | 0.123 | 0.177 | 0.787 | 0.123 |
| *Spina Bifida* | 0.567 | 0.450 | 0.450 | 0.527 | 0.450 | 0.450 | 0.450 | 0.641 | - | 0.798 | - | - | - | 0.276 | 0.798 | - | - | - | 0.276 | 0.798 | 0.798 | 0.798 | 0.316 | - | - | 0.276 | - | 0.276 | 0.276 |
| *Cerebral Palsy* | 0.247 | 0.478 | 0.392 | 0.392 | 0.392 | 0.495 | 0.392 | 0.392 | 0.765 | 0.765 | 0.765 | 0.210 | 0.765 | 0.765 | - | - | 0.177 | - | - | - | 0.177 | - | - | 0.177 | - | - | 0.177 | 0.177 | - |
| **Injury, Poisoning & Procedural Complications:** | | | | | | | | | | | | | | | | | | | | | | | | | | | | | |
| Total ADRs: | <0.05 | <0.05 | <0.05 | <0.05 | <0.05 | 0.714 | <0.05 | <0.05 | <0.05 | 0.123 | 0.508 | <0.05 | 0.736 | 0.423 | 0.259 | <0.05 | <0.05 | <0.05 | 0.064 | 0.370 | <0.05 | 0.224 | 0.449 | <0.05 | 0.744 | 0.889 | <0.05 | <0.05 | 0.641 |
| Fatalities | 0.958 | 0.884 | 0.685 | 0.707 | 0.687 | 0.687 | 0.687 | 0.687 | 0.599 | 0.618 | 0.765 | 0.765 | 0.765 | 0.765 | 0.975 | 0.484 | 0.484 | 0.484 | 0.484 | 0.484 | 0.484 | 0.484 | 0.484 | - | - | - | - | - | - |
| Exposures, chemical injuries, and poisoning | <0.05 | <0.05 | <0.05 | <0.05 | <0.05 | <0.05 | <0.05 | <0.05 | 0.364 | 0.767 | 0.783 | 0.742 | 0.767 | 0.492 | 0.239 | 0.247 | 0.549 | 0.239 | 0.809 | 0.984 | 0.535 | 1.000 | 0.334 | 0.548 | 0.984 | 0.344 | 0.535 | 0.714 | 0.334 |
| *Exposure in Pregnancy* | 0.556 | 0.344 | 0.321 | 0.503 | 0.392 | 0.691 | 0.552 | 0.286 | 0.926 | 0.712 | 0.885 | 0.210 | 0.656 | 0.765 | 0.658 | 0.817 | 0.197 | 0.607 | 0.815 | 0.813 | 0.308 | 0.931 | 0.567 | 0.238 | 0.750 | 0.687 | 0.341 | 0.177 | 0.526 |
| *Foetal exposure during pregnancy* | <0.05 | 0.002 | 0.001 | 0.007 | 0.007 | 0.001 | <0.05 | <0.05 | 0.625 | 0.520 | 0.543 | 0.463 | 0.313 | 0.620 | 0.298 | 0.313 | 0.712 | 0.176 | 0.361 | 0.969 | 0.229 | 0.688 | 0.875 | 0.240 | 0.661 | 0.905 | 0.137 | 0.276 | 0.579 |
| *Maternal Exposure in Pregnancy* | 0.908 | 0.523 | 0.564 | 0.903 | 0.670 | 0.523 | 0.845 | 0.523 | 0.869 | 0.469 | 0.345 | - | 0.437 | - | 0.503 | 0.367 | 0.869 | 0.468 | 0.869 | 0.758 | 0.469 | 0.942 | 0.469 | 0.345 | 0.813 | 0.345 | 0.437 | - | 0.437 |
| *Toxicity to various agents* | 0.399 | 0.786 | 0.154 | 0.429 | 0.330 | 0.154 | 0.154 | 0.154 | 0.217 | 0.593 | 0.465 | 0.217 | 0.217 | 0.217 | 0.395 | 0.485 | - | - | - | 0.831 | 0.395 | 0.395 | 0.395 | 0.485 | 0.485 | 0.485 | - | - | - |
| Off Label Uses | 0.058 | 0.205 | 0.085 | 0.090 | 0.755 | 0.312 | 0.673 | 0.542 | 0.547 | 0.574 | 0.325 | 0.032 | 0.378 | 0.483 | 0.964 | 0.141 | <0.05 | 0.167 | 0.221 | 0.149 | <0.05 | 0.177 | 0.233 | 0.191 | 0.912 | 0.762 | 0.159 | 0.114 | 0.848 |
| **Pregnancy, Puerperium & Perinatal Conditions:** | | | | | | | | | | | | | | | | | | | | | | | | | | | | | |
| Total ADRs: | 0.655 | 0.333 | 0.138 | 0.693 | 0.404 | 0.822 | 0.323 | 0.131 | 0.458 | 0.546 | 0.878 | 0.445 | 0.983 | 0.428 | 0.232 | 0.392 | 0.187 | 0.468 | 0.869 | 0.647 | 0.864 | 0.533 | 0.219 | 0.533 | 0.861 | 0.368 | 0.433 | 0.177 | 0.437 |
| Fatalities | - | - | - | - | - | - | - | - | - | - | - | - | - | - | - | - | - | - | - | - | - | - | - | - | - | - | - | - | - |
| Abortions and Still Births | 0.667 | 0.429 | 0.304 | 0.489 | 0.304 | 0.304 | 0.304 | 0.304 | 0.672 | 0.900 | 0.672 | 0.672 | 0.672 | 0.672 | 0.608 | - | - | - | - | 0.608 | 0.608 | 0.608 | 0.608 | - | - | - | - | - | - |
| Foetal Complications | 0.998 | 0.687 | 0.687 | 0.954 | 0.999 | 0.687 | 0.687 | 0.687 | - | 0.717 | 0.687 | - | - | - | 0.717 | 0.687 | - | - | - | 0.954 | 0.717 | 0.717 | 0.717 | 0.687 | 0.687 | 0.687 | - | - | - |
| Neonatal and Perinatal Conditions | 0.977 | 0.789 | 0.677 | 0.943 | 0.898 | 0.621 | 0.697 | 0.621 | 0.855 | 0.841 | 0.885 | 0.765 | 0.537 | 0.765 | 0.720 | 0.756 | 0.869 | 0.468 | 0.869 | 0.954 | 0.657 | 0.650 | 0.657 | 0.687 | 0.614 | 0.687 | 0.437 | - | 0.437 |
| Pregnancy, Labour, Delivery and | 0.474 | 0.676 | 0.775 | 0.449 | 0.703 | 0.207 | 0.775 | 0.775 | 0.549 | 0.688 | 0.967 | 0.321 | 0.549 | 0.549 | 0.375 | 0.569 | 0.177 | - | - | 0.661 | 0.522 | 0.375 | 0.375 | 0.306 | 0.569 | 0.569 | 0.177 | 0.177 | - |

| | | | | | | | | | | | | | | | | | | | | | | | | | | | | | |
|---|---|---|---|---|---|---|---|---|---|---|---|---|---|---|---|---|---|---|---|---|---|---|---|---|---|---|---|---|---|
| Postpartum Conditions | | | | | | | | | | | | | | | | | | | | | | | | | | | | | |
| **OTHER SIGNIFICANT ADRs:** | | | | | | | | | | | | | | | | | | | | | | | | | | | | | |
| **Gastrointestinal Disorders:** | | | | | | | | | | | | | | | | | | | | | | | | | | | | | |
| Total ADRs: | <0.05 | 0.612 | 0.141 | 0.162 | 0.142 | 0.252 | 0.263 | 0.273 | 0.328 | 0.366 | 0.329 | 0.101 | 0.106 | 0.552 | 0.941 | 0.999 | <0.05 | <0.05 | 0.698 | 0.942 | <0.05 | <0.05 | 0.754 | <0.05 | <0.05 | 0.699 | 0.980 | <0.05 | <0.05 |
| Fatalities | 0.997 | 0.687 | 0.687 | 0.687 | 0.869 | 0.687 | 0.687 | 0.687 | - | - | 0.776 | - | - | - | - | 0.776 | - | - | - | 0.776 | - | - | - | 0.776 | 0.776 | 0.776 | - | - | - |
| Exocrine Pancreas Conditions | 0.120 | 0.118 | 0.137 | 0.167 | 0.180 | 0.118 | 0.226 | 0.118 | 0.775 | 0.657 | 0.622 | - | 0.526 | - | 0.827 | 0.777 | 0.775 | 0.645 | 0.775 | 0.944 | 0.657 | 0.791 | 0.657 | 0.622 | 0.844 | 0.622 | 0.526 | - | 0.526 |
| *Diarrhoea* | 0.576 | 0.842 | 0.854 | 0.619 | 0.902 | 0.613 | 0.489 | 0.224 | 0.987 | 0.759 | 0.939 | 0.487 | 0.382 | 0.169 | 0.748 | 0.952 | 0.497 | 0.390 | 0.172 | 0.704 | 0.337 | 0.260 | 0.111 | 0.533 | 0.421 | 0.188 | 0.847 | 0.453 | 0.573 |
| *Constipation* | 0.958 | 0.904 | 0.664 | 0.736 | 0.545 | 0.450 | 0.842 | 0.450 | 0.748 | 0.826 | 0.614 | 0.503 | 0.750 | 0.503 | 0.915 | 0.833 | 0.662 | 0.539 | 0.662 | 0.757 | 0.608 | 0.600 | 0.608 | 0.776 | 0.442 | 0.776 | 0.370 | - | 0.370 |
| *Nausea and Vomiting* | <0.05 | 0.824 | 0.911 | 0.994 | 0.790 | <0.05 | 0.149 | 0.493 | 0.911 | 0.830 | 0.627 | <0.05 | 0.214 | 0.639 | 0.918 | 0.706 | <0.05 | 0.179 | 0.563 | 0.784 | <0.05 | 0.151 | 0.497 | <0.05 | 0.095 | 0.348 | 0.059 | <0.05 | 0.426 |
| **General Disorders and Administration Site Conditions** | | | | | | | | | | | | | | | | | | | | | | | | | | | | | |
| Total ADRs | <0.05 | 0.147 | <0.05 | <0.05 | <0.05 | <0.05 | 0.224 | <0.05 | <0.05 | 0.245 | 0.552 | <0.05 | 0.811 | <0.05 | 0.382 | 0.151 | <0.05 | <0.05 | <0.05 | 0.568 | <0.05 | 0.162 | <0.05 | <0.05 | 0.405 | <0.05 | <0.05 | 0.236 | <0.05 |
| Fatalities | 0.443 | 0.771 | 0.616 | 0.622 | 0.643 | 0.420 | 0.626 | 0.321 | 0.821 | 0.828 | 0.465 | 0.549 | 0.833 | 0.222 | 0.992 | 0.364 | 0.662 | 0.987 | 0.172 | 0.368 | 0.657 | 0.995 | 0.174 | 0.255 | 0.370 | 0.574 | 0.654 | 0.123 | 0.175 |
| General Systems Disorders NEC | <0.05 | 0.186 | <0.05 | <0.05 | <0.05 | 0.325 | 0.482 | 0.366 | 0.206 | 0.394 | 0.329 | <0.05 | 0.531 | <0.05 | 0.674 | 0.768 | <0.05 | 0.062 | <0.05 | 0.900 | <0.05 | 0.142 | <0.05 | <0.05 | 0.112 | <0.05 | 0.094 | 0.936 | 0.110 |
| *Developmental Delay* | 0.194 | 0.227 | 0.191 | 0.191 | 0.258 | 0.191 | 0.191 | 0.191 | 0.765 | 0.765 | 0.885 | 0.765 | 0.765 | 0.765 | - | 0.687 | - | - | - | 0.687 | - | - | - | 0.687 | 0.687 | 0.687 | - | - | - |
| *Fatigue* | 0.066 | 0.970 | 0.459 | 0.772 | 0.830 | 0.053 | 0.544 | 0.108 | 0.439 | 0.800 | 0.859 | 0.057 | 0.568 | 0.115 | 0.317 | 0.351 | <0.05 | 0.201 | <0.05 | 0.939 | 0.090 | 0.748 | 0.175 | 0.079 | 0.691 | 0.155 | 0.159 | 0.710 | 0.290 |
| *Drug Interaction* | <0.05 | 0.546 | 0.142 | 0.458 | 0.196 | 0.182 | 0.205 | 0.083 | 0.334 | 0.884 | 0.452 | 0.062 | 0.469 | 0.192 | 0.401 | 0.799 | <0.05 | 0.775 | 0.567 | 0.537 | <0.05 | 0.557 | 0.229 | <0.05 | 0.974 | 0.451 | <0.05 | <0.05 | 0.437 |
| *Drug Ineffective* | <0.05 | 0.712 | 0.168 | 0.270 | 0.774 | <0.05 | 0.709 | <0.05 | 0.296 | 0.451 | 0.513 | <0.05 | 0.997 | <0.05 | 0.752 | 0.103 | <0.05 | 0.298 | <0.05 | 0.171 | <0.05 | 0.454 | <0.05 | <0.05 | 0.511 | 0.062 | <0.05 | 0.356 | <0.05 |
| **Nervous System Disorders:** | | | | | | | | | | | | | | | | | | | | | | | | | | | | | |
| Total ADRs: | <0.05 | <0.05 | <0.05 | <0.05 | <0.05 | <0.05 | 0.889 | 0.857 | <0.05 | 0.250 | 0.904 | <0.05 | <0.05 | <0.05 | 0.118 | <0.05 | <0.05 | <0.05 | <0.05 | 0.204 | <0.05 | <0.05 | <0.05 | <0.05 | 0.056 | <0.05 | <0.05 | <0.05 | 0.749 |
| Fatalities: | 0.999 | 0.884 | 0.756 | 0.687 | 0.869 | 0.687 | 0.687 | 0.687 | 0.855 | 0.765 | 0.984 | 0.765 | 0.765 | 0.765 | 0.869 | 0.870 | 0.869 | 0.869 | 0.869 | 0.776 | - | - | - | 0.776 | 0.776 | 0.776 | - | - | - |
| Neurological Disorders NEC | <0.05 | 0.805 | 0.067 | 0.168 | 0.246 | 0.086 | 0.097 | 0.194 | 0.110 | 0.254 | 0.359 | 0.051 | 0.058 | 0.123 | 0.633 | 0.483 | <0.05 | <0.05 | <0.05 | 0.821 | <0.05 | <0.05 | <0.05 | <0.05 | <0.05 | <0.05 | 0.955 | 0.671 | 0.713 |
| *Speech Disorder* | 0.916 | 0.721 | 0.467 | 0.472 | 0.445 | 0.323 | 0.897 | 0.886 | 0.685 | 0.691 | 0.654 | 0.463 | 0.818 | 0.620 | 0.992 | 0.962 | 0.662 | 0.539 | 0.396 | 0.954 | 0.657 | 0.544 | 0.400 | 0.687 | 0.514 | 0.377 | 0.370 | 0.276 | 0.785 |
| *Speech Disorder Developmental* | 0.503 | 0.366 | 0.304 | 0.349 | 0.304 | 0.304 | 0.304 | 0.304 | 0.765 | 0.952 | 0.765 | 0.765 | 0.765 | 0.765 | 0.798 | - | - | - | - | 0.798 | 0.798 | 0.798 | 0.798 | - | - | - | - | - | - |
| *Dyspraxia* | 0.433 | 0.286 | 0.286 | 0.371 | 0.286 | 0.286 | 0.286 | 0.286 | - | 0.717 | - | - | - | - | 0.717 | - | - | - | - | 0.717 | 0.717 | 0.717 | 0.717 | - | - | - | - | - | - |
| *Dyslexia* | 0.987 | 0.941 | 0.568 | 0.678 | 0.702 | 0.568 | 0.568 | 0.568 | 0.604 | 0.725 | 0.751 | 0.604 | 0.604 | 0.604 | 0.798 | 0.776 | - | - | - | 0.968 | 0.798 | 0.798 | 0.798 | 0.776 | 0.776 | 0.776 | - | - | - |
| *Epilepsy* | <0.05 | 0.719 | 0.091 | 0.400 | 0.560 | 0.090 | 0.143 | 0.880 | 0.155 | 0.621 | 0.821 | <0.05 | 0.244 | 0.611 | 0.298 | 0.211 | <0.05 | 0.682 | 0.071 | 0.787 | <0.05 | 0.464 | 0.325 | <0.05 | 0.331 | 0.466 | <0.05 | 0.119 | 0.112 |
| Seizures | <0.05 | 0.548 | <0.05 | 0.183 | 0.800 | 0.000 | 0.369 | <0.05 | <0.05 | 0.450 | 0.727 | <0.05 | 0.762 | 0.131 | 0.173 | <0.05 | <0.05 | 0.083 | 0.520 | 0.274 | <0.05 | 0.647 | 0.415 | <0.05 | 0.516 | 0.069 | <0.05 | <0.05 | 0.215 |
| Mental Impairment Disorder | 0.277 | 0.204 | 0.193 | 0.467 | 0.486 | 0.367 | 0.896 | 0.862 | 0.972 | 0.560 | 0.539 | <0.05 | 0.165 | 0.153 | 0.537 | 0.517 | <0.05 | 0.156 | 0.145 | 0.974 | 0.113 | 0.393 | 0.370 | 0.119 | 0.410 | 0.387 | 0.439 | 0.464 | 0.966 |
| Movement Disorders | <0.05 | 0.086 | 0.065 | 0.088 | 0.080 | 0.289 | 0.197 | 0.166 | 0.872 | 0.987 | 0.968 | <0.05 | 0.627 | 0.702 | 0.859 | 0.903 | <0.05 | 0.520 | 0.589 | 0.956 | <0.05 | 0.637 | 0.713 | <0.05 | 0.599 | 0.673 | <0.05 | <0.05 | 0.916 |
| **Psychiatric Disorders:** | | | | | | | | | | | | | | | | | | | | | | | | | | | | | |
| Total ADRs: | <0.05 | <0.05 | <0.05 | <0.05 | 0.597 | <0.05 | 0.767 | 0.830 | 0.397 | 0.920 | 0.114 | <0.05 | <0.05 | <0.05 | 0.344 | <0.05 | <0.05 | <0.05 | <0.05 | 0.138 | <0.05 | <0.05 | <0.05 | <0.05 | 0.410 | 0.458 | <0.05 | <0.05 | 0.935 |
| Fatalities: | 0.998 | 0.789 | 0.729 | 0.943 | 0.898 | 0.621 | 0.621 | 0.621 | 0.926 | 0.841 | 0.885 | 0.765 | 0.765 | 0.765 | 0.776 | 0.817 | 0.815 | 0.815 | 0.815 | 0.954 | 0.657 | 0.657 | 0.657 | 0.687 | 0.687 | 0.687 | - | - | - |
| Anxiety Disorders and Symptoms | <0.05 | 0.947 | 0.635 | 0.987 | 0.631 | <0.05 | 0.387 | 0.332 | 0.682 | 0.934 | 0.585 | <0.05 | 0.354 | 0.302 | 0.623 | 0.348 | <0.05 | 0.193 | 0.162 | 0.642 | <0.05 | 0.396 | 0.340 | <0.05 | 0.694 | 0.616 | 0.096 | 0.118 | 0.913 |
| Cognitive and attention disorders | <0.05 | 0.121 | <0.05 | 0.064 | 0.050 | <0.05 | 0.053 | 0.183 | 0.418 | 0.654 | 0.513 | 0.369 | 0.546 | 0.787 | 0.658 | 0.817 | 0.815 | 0.772 | 0.309 | 0.813 | 0.567 | 0.861 | 0.485 | 0.687 | 0.949 | 0.377 | 0.654 | 0.276 | 0.402 |
| *Learning Disorders* | 0.062 | 0.163 | 0.071 | 0.071 | 0.082 | 0.071 | 0.100 | 0.327 | 0.463 | 0.463 | 0.562 | 0.463 | 0.695 | 0.620 | - | 0.776 | - | 0.654 | 0.276 | 0.776 | - | 0.654 | 0.276 | 0.776 | 0.821 | 0.326 | 0.654 | 0.276 | 0.402 |
| *ADHD* | 0.789 | 0.458 | 0.343 | 0.570 | 0.384 | 0.323 | 0.323 | 0.323 | 0.738 | 0.834 | 0.848 | 0.672 | 0.672 | 0.672 | 0.614 | 0.870 | 0.869 | 0.869 | 0.869 | 0.699 | 0.567 | 0.567 | 0.567 | 0.776 | 0.776 | 0.776 | - | - | - |
| *Autism Spectrum Disorders* | <0.05 | 0.213 | 0.075 | 0.116 | 0.128 | 0.075 | 0.075 | 0.075 | 0.397 | 0.646 | 0.701 | 0.397 | 0.397 | 0.397 | 0.608 | 0.569 | - | - | - | 0.935 | 0.608 | 0.608 | 0.608 | 0.569 | 0.569 | 0.569 | - | - | - |
| *Neurodevelopmental Disorders* | <0.05 | 0.687 | 0.687 | 0.954 | 0.687 | 0.687 | 0.687 | 0.687 | - | 0.717 | - | - | - | - | 0.717 | - | - | - | - | 0.717 | 0.717 | 0.717 | 0.717 | - | - | - | - | - | - |
| Deliria | <0.05 | 0.752 | 0.472 | 0.456 | 0.652 | 0.120 | 0.806 | 0.118 | 0.679 | 0.660 | 0.891 | 0.068 | 0.577 | 0.181 | 0.978 | 0.781 | <0.05 | 0.342 | 0.296 | 0.760 | <0.05 | 0.330 | 0.305 | 0.053 | 0.490 | 0.214 | 0.183 | <0.05 | 0.082 |
| **Respiratory, Thoracic and Mediastinal Disorders:** | | | | | | | | | | | | | | | | | | | | | | | | | | | | | |
| Total ADRs: | <0.05 | 0.929 | 0.151 | 0.351 | 0.282 | 0.158 | 0.708 | 0.140 | 0.129 | 0.308 | 0.245 | 0.135 | 0.775 | 0.164 | 0.586 | 0.693 | 0.978 | 0.076 | <0.05 | 0.879 | 0.605 | 0.197 | <0.05 | 0.713 | 0.153 | <0.05 | 0.080 | <0.05 | 0.265 |
| Fatalities: | 0.999 | 0.775 | 0.999 | 0.775 | 0.775 | 0.775 | 0.775 | 0.775 | 0.775 | - | - | - | - | - | 0.775 | 0.775 | 0.775 | 0.775 | 0.775 | - | - | - | - | - | - | - | - | - | - |
| *Breathing Abnormalities* | 0.139 | 0.572 | 0.445 | 0.542 | 0.492 | 0.947 | 0.977 | 0.156 | 0.832 | 0.962 | 0.897 | 0.617 | 0.554 | 0.060 | 0.869 | 0.934 | 0.483 | 0.430 | <0.05 | 0.935 | 0.585 | 0.524 | 0.055 | 0.533 | 0.476 | <0.05 | 0.924 | 0.139 | 0.163 |
| **Skin and Subcutaneous Tissue Disorders:** | | | | | | | | | | | | | | | | | | | | | | | | | | | | | |
| Total ADRs: | <0.05 | <0.05 | 0.348 | <0.05 | 0.728 | <0.05 | 0.309 | <0.05 | <0.05 | 0.831 | <0.05 | 0.899 | 0.085 | 0.097 | <0.05 | 0.203 | <0.05 | 0.056 | <0.05 | <0.05 | 0.734 | 0.129 | 0.062 | <0.05 | 0.500 | <0.05 | 0.065 | 0.125 | <0.05 |
| Fatalities: | - | - | - | - | - | - | - | - | - | - | - | - | - | - | - | - | - | - | - | - | - | - | - | - | - | - | - | - | - |
| Rashes, eruptions and exanthems NEC | <0.05 | <0.05 | 0.742 | 0.054 | 0.859 | <0.05 | 0.973 | 0.187 | <0.05 | 0.873 | 0.055 | 0.453 | <0.05 | 0.403 | <0.05 | 0.615 | <0.05 | 0.768 | 0.112 | 0.075 | 0.364 | 0.051 | 0.497 | <0.05 | 0.832 | 0.246 | <0.05 | 0.118 | 0.177 |

**Table S4.** Physiochemical and pharmacokinetic properties of the eight AEDs studied.

| | Valproic acid | Carbamazepine | Gabapentin | Lamotrigine | Levetiracetam | Oxcarbazepine | Topiramate | Zonisamide |
|---|---|---|---|---|---|---|---|---|
| **Physiochemical:** | | | | | | | | |
| MW (Da) | 144.21 | 236.27 | 171.24 | 256.1 | 170.21 | 252.27 | 339.37 | 212.23 |
| p$K_a$ | 5.14 | - | 4.63 | 5.89 | - | - | 11.09 | - |
| $cLog_{10}P$ | 2.8 | 2.77 | -1.27 | 1.93 | -0.59 | 1.82 | 0.13 | -0.03 |
| $cLog_{10}D_{7.4}$ | 0.56 | 2.77 | -1.27 | 1.91 | -0.59 | 1.82 | 0.13 | -1.68 |
| $^{t}$PSA (Å) | 37.3 | 46.33 | 63.32 | 90.71 | 63.4 | 63.4 | 115.54 | 87.18 |
| HBA | 1 | 1 | 2 | 5 | 2 | 2 | 8 | 4 |
| HBD | 1 | 1 | 2 | 2 | 1 | 1 | 1 | 2 |
| LLE | 1.6 | 1.9 | 8.8 | 2.3 | 3.2 | 2.7 | 6.2 | 7.5 |
| p-glycoprotein substrate | No | Yes | No | Yes | Yes | Yes | Yes | No |
| $pIC_{50}$ | 4.4 | 4.7 | 7.6 | 4.2 | 2.7 | 4.6 | 6.3 | 7.5 |
| **Pharmacokinetics:** | | | | | | | | |
| $t_{1/2}$ (h) | 8-20 | 36 | 5-7 | 33 | 7 | Ox: 1.3-2.3, MHD: 9.3 | 21 | 60 |
| $T_{max}$ (h) | 1.72 | 24 | 2.7 | 2.5 | 1.3 | 4.5 | 2-3 | 2-5 |
| $C_{max}$ (nM) | 184315 | 11004-14814 | 23476 | 17962 | 123377 | 34000 | 4420 | 9424 |

| | | | | | | | | |
|---|---|---|---|---|---|---|---|---|
| %*F* (%) | Approx. 100 | 70-78 | 60 | 98 | Approx. 100 | >95 | 81 | Approx. 100 |
| $V_d$ (L/kg) | 0.1-0.4 | 1.9 | 0.8 | 0.92-1.22 | 0.5-0.7 | 0.8 | 0.8 | 1.7 |
| Renal Elimination (%) | 30-50 | 72 | 100 | 94 | 66 | 95 | 70-80 | 62 |
| PPB (%) | 80-95 | 75 | <3 | 55 | <10 | MHD: 40 | 13.10 | 40-50 |
| Clearance (L/h) | 0.3-0.6 (P) | 1.5 (O) | 5.4-6 (R) | 1.8 (P), 2.5 (O) | 0.96 mL/kg/min (P), 0.6 mL/kg/min (R) | Ox: 84.9 MHD: 2.0 | 1.02 (R) 1.2-1.8 (P) | 0.21 (R) |
| Dosing | OD / BD | OD / BD | TDS | OD / BD | OD / BD | BD | BD | OD |
| Dose Used for $C_{max}$ | 250mg Tablet | 200mg Tablet | 300mg Tablet | 400mg Tablet | 750mg Tablet | 600mg Tablet | 100mg Tablet | 200mg Tablet |

(OD) = Once Daily, (BD) = Twice Daily, (TDS) = Three times daily, $cLog_{10}P$ = Calculated $Log_{10}P$, $cLog_{10}D_{7.4}$ = Calculated $Log_{10}D$ at pH 7.4, HBA = Hydrogen bond acceptor, HBD = Hydrogen bond donor, (P) = Plasma, (R) = Renal, MHD = 10-Monohydroxy derivative (oxcarbazepine metabolite), $C_{max}$ = Peak Serum concentration, $^{t}$PSA = Total polar surface area, %F = bioavailability, PPB = plasma protein binding, $t_{max}$ = time taken to reach $C_{max}$, PPB = Plasma Protein binding, LLE = Ligand Lipophilic Efficiency, $pK_a$ = Acid dissociation constant, MW = Molecular Weight, $V_d$= Volume of distribution, BBB = blood brain barrier, $pIC_{50}$ calculated for main protein target.

**Chart S1:** Number of Prescriptions dispensed per AED between August 2017 – July 2022

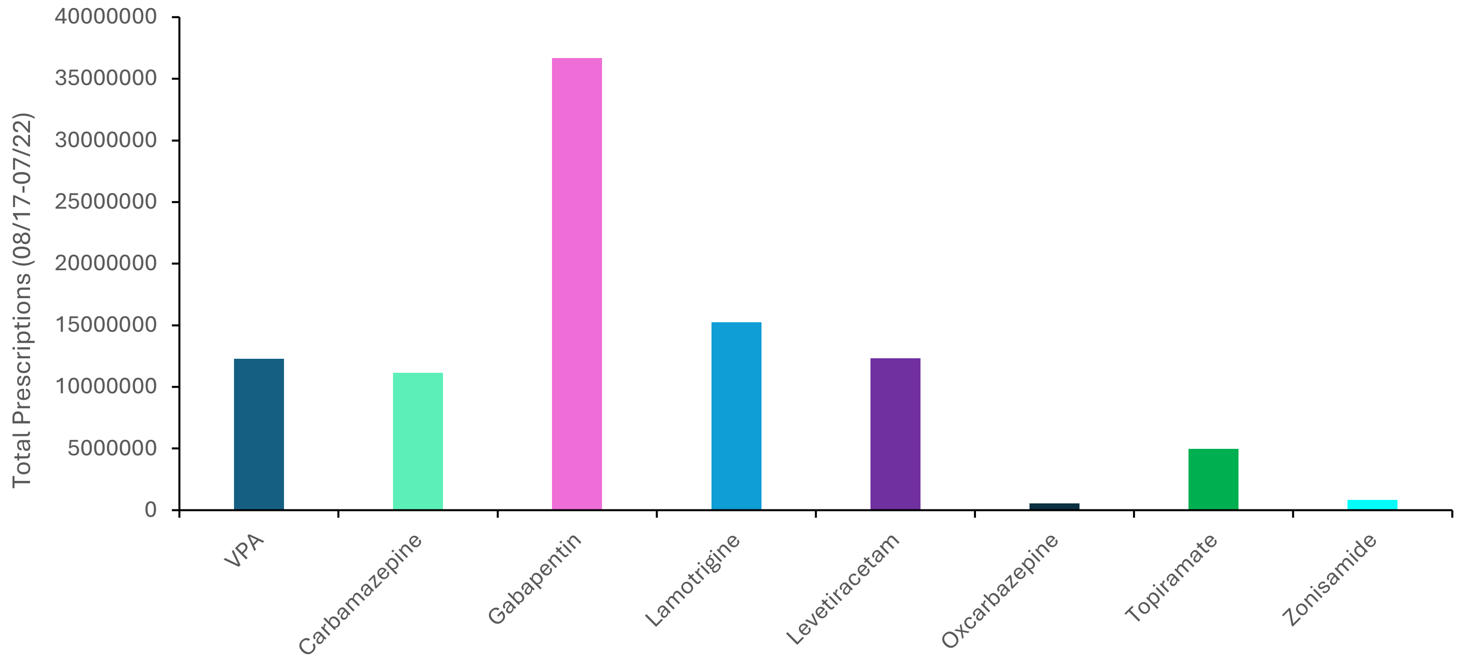

**Chart S2:** Number of standardised reported AEDs per 1 million prescriptions between January 2018-August 2022:

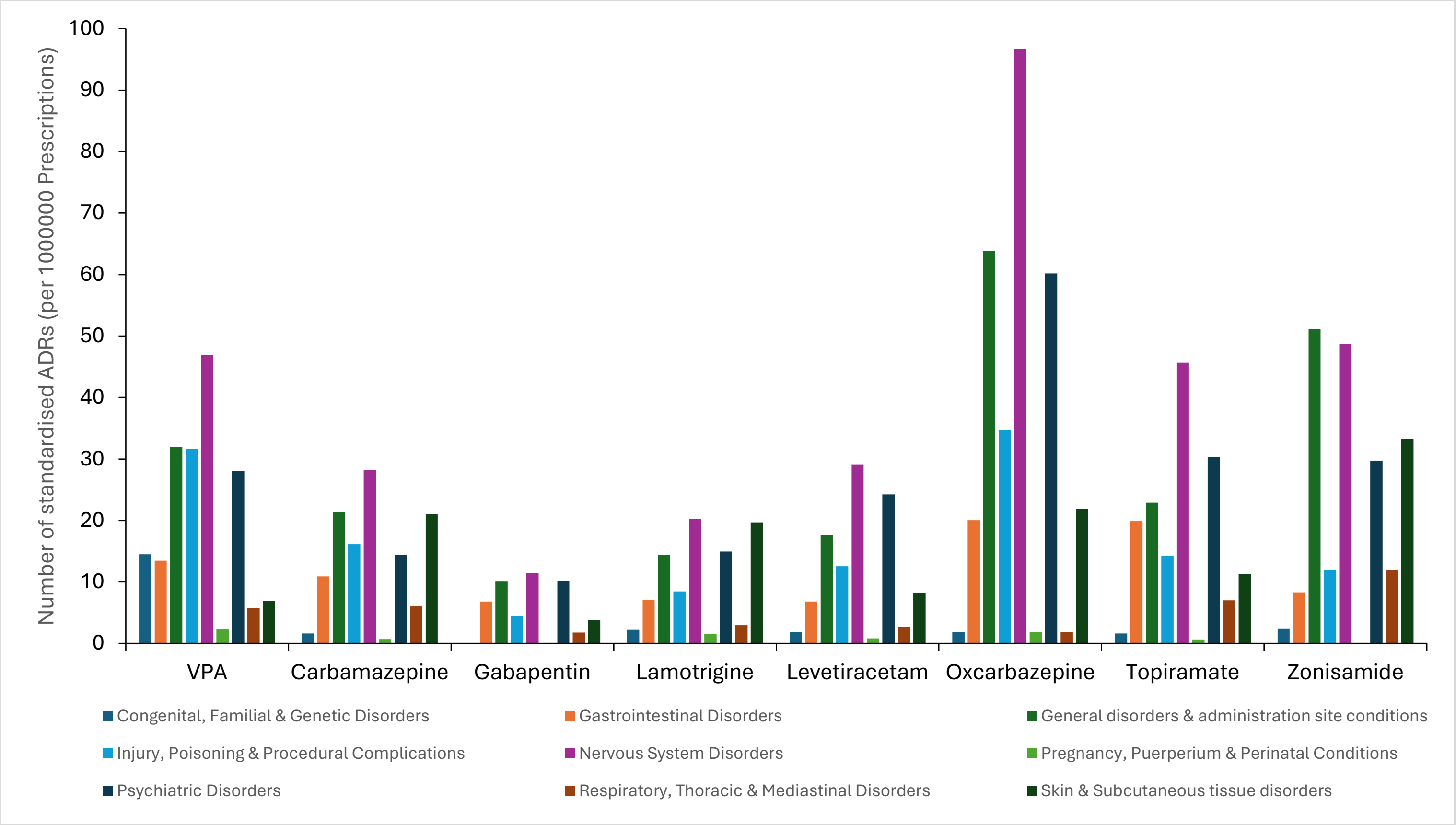

**Chart S3:** Total number of standardised reported suspected fatalities per 1 million prescriptions between January 2018-August 2022:

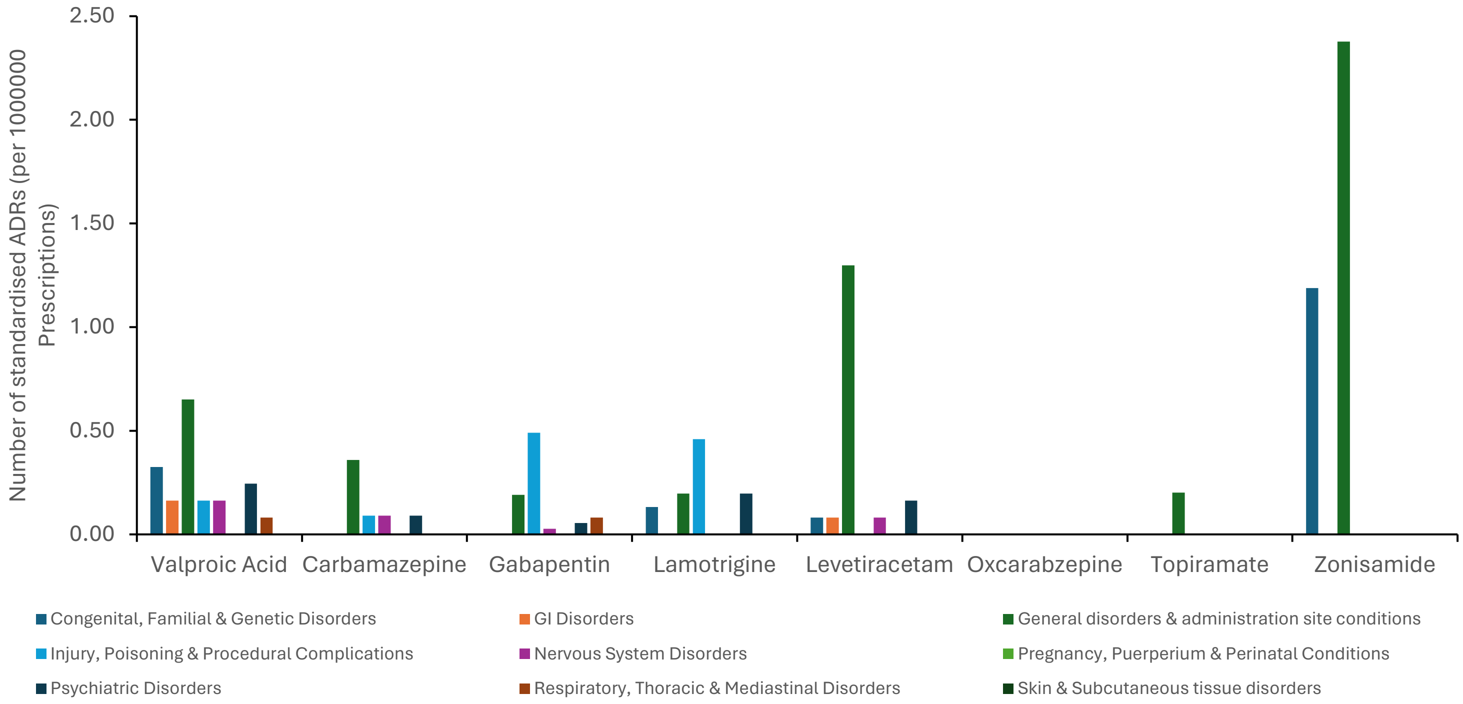

**Figure S2.** Odds ratio and confidence interval for VPA.

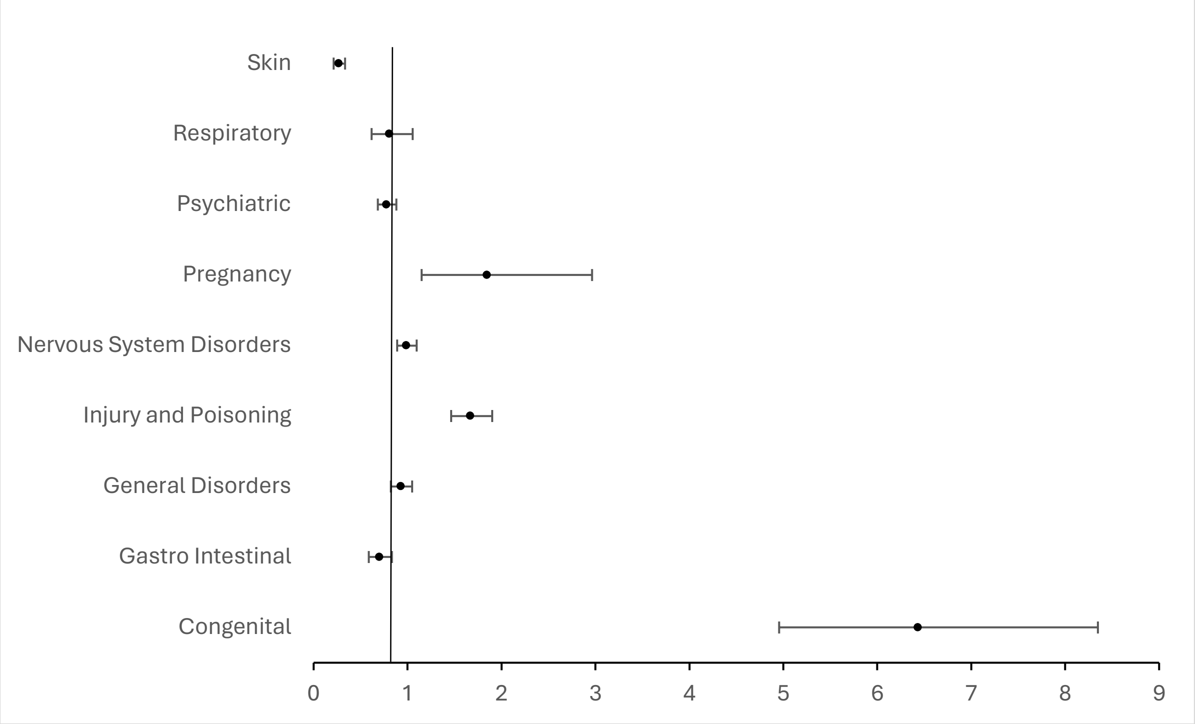

**Figure S3.** Odds ratio and confidence interval for carbamazepine.

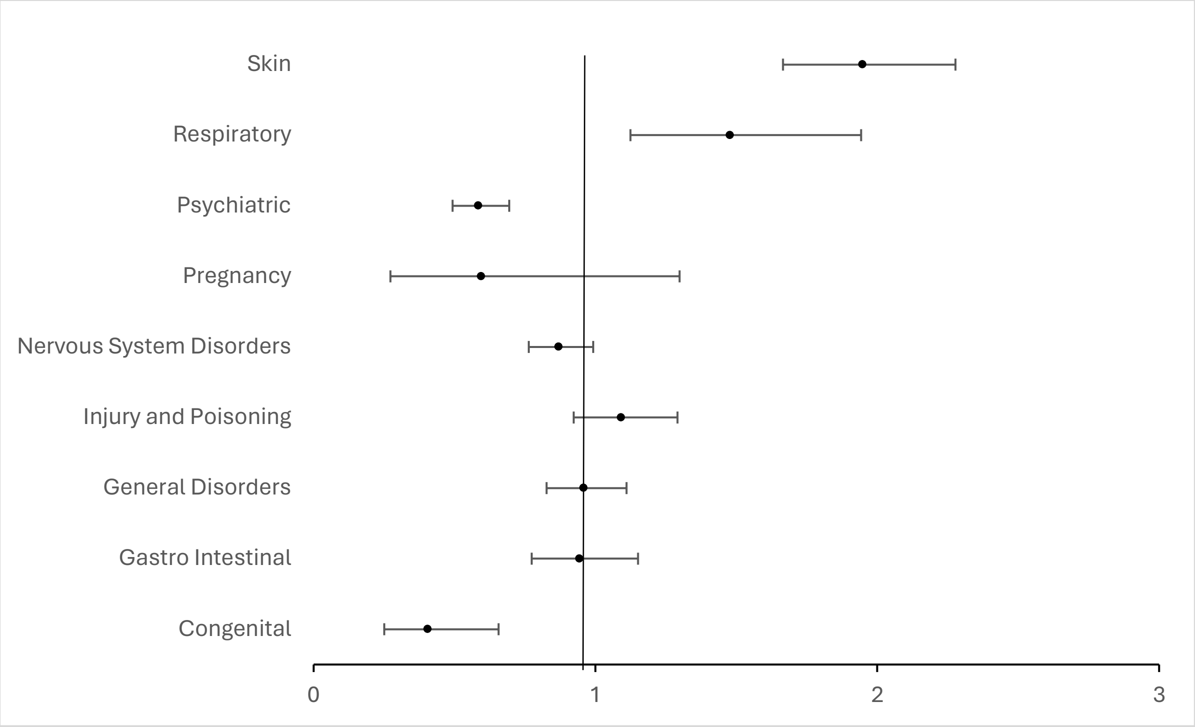

**Figure S4.** Odds ratio and confidence interval for gabapentin.

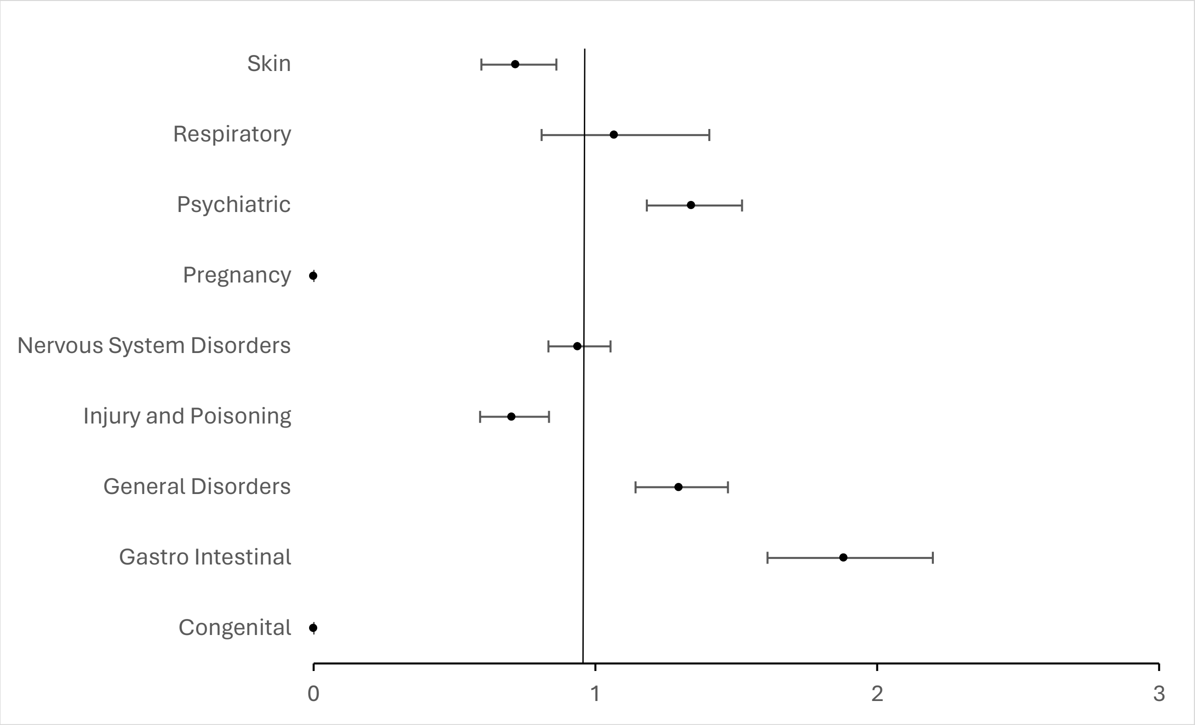

**Figure S5.** Odds ratio and confidence interval for lamotrigine.

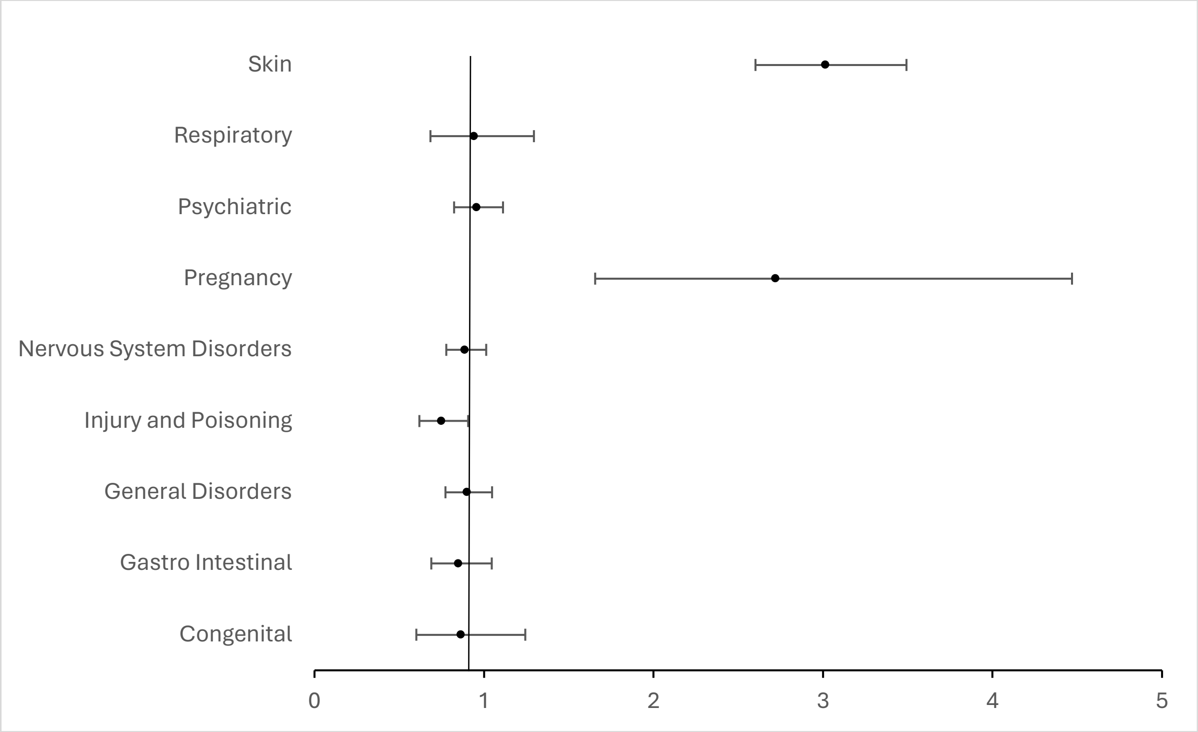

**Figure S6.** Odds ratio and confidence interval for levetiracetam.

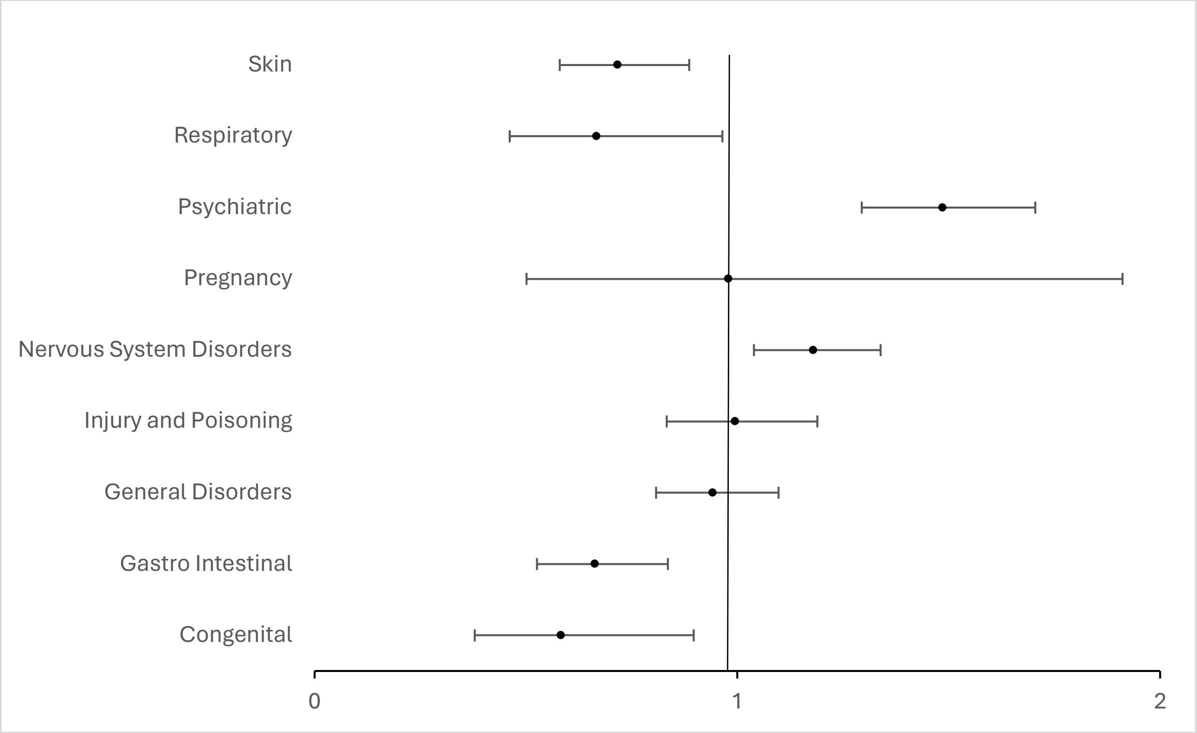

**Figure S7.** Odds ratio and confidence interval for oxcarbazepine.

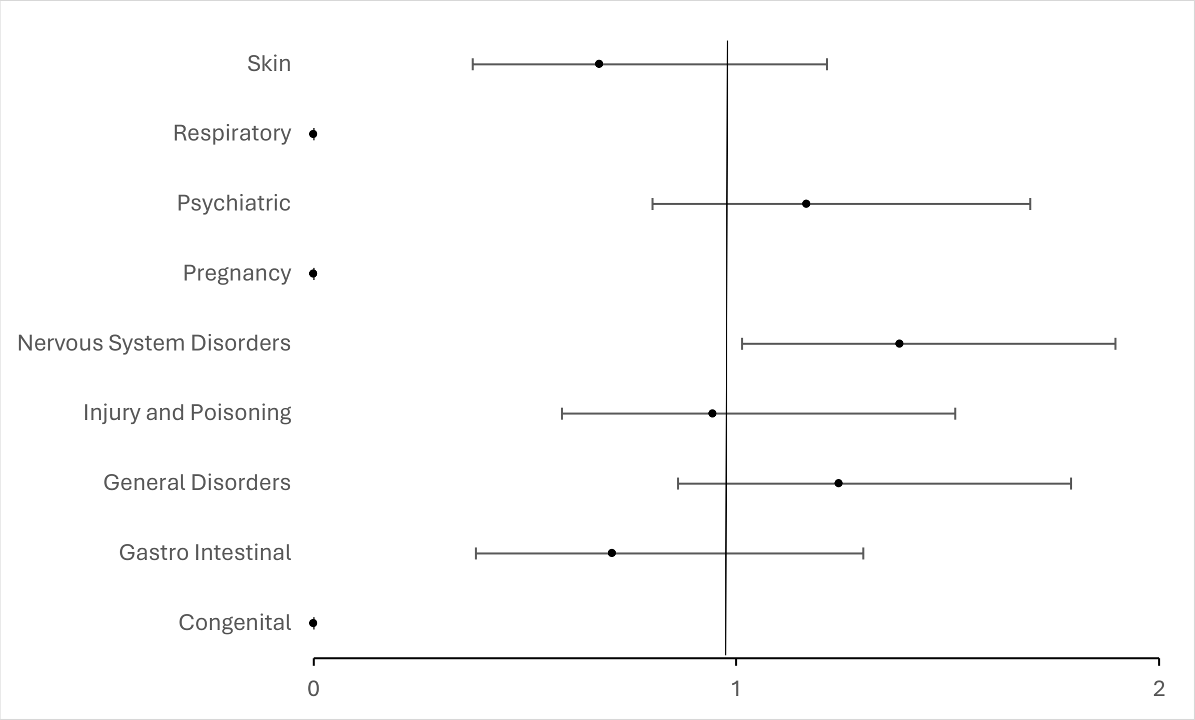

**Figure S8.** Odds ratio and confidence interval for topiramate.

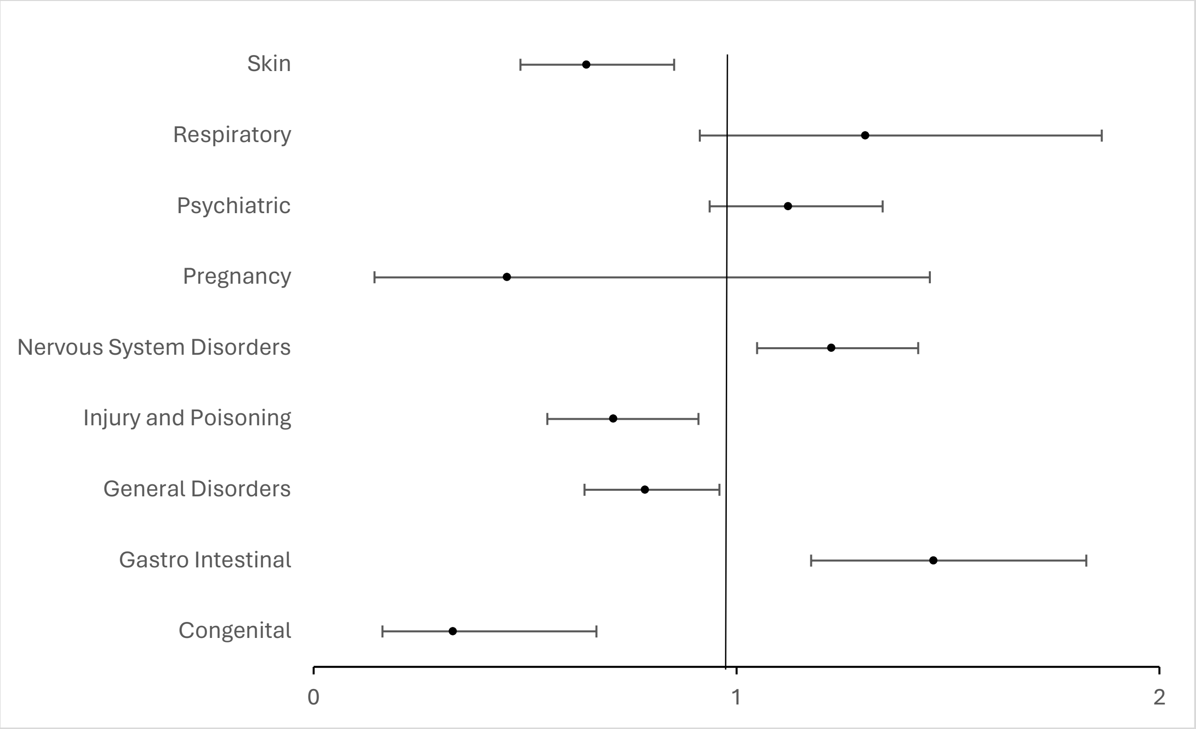

**Figure S9.** Odds ratio and confidence interval for zonisamide.

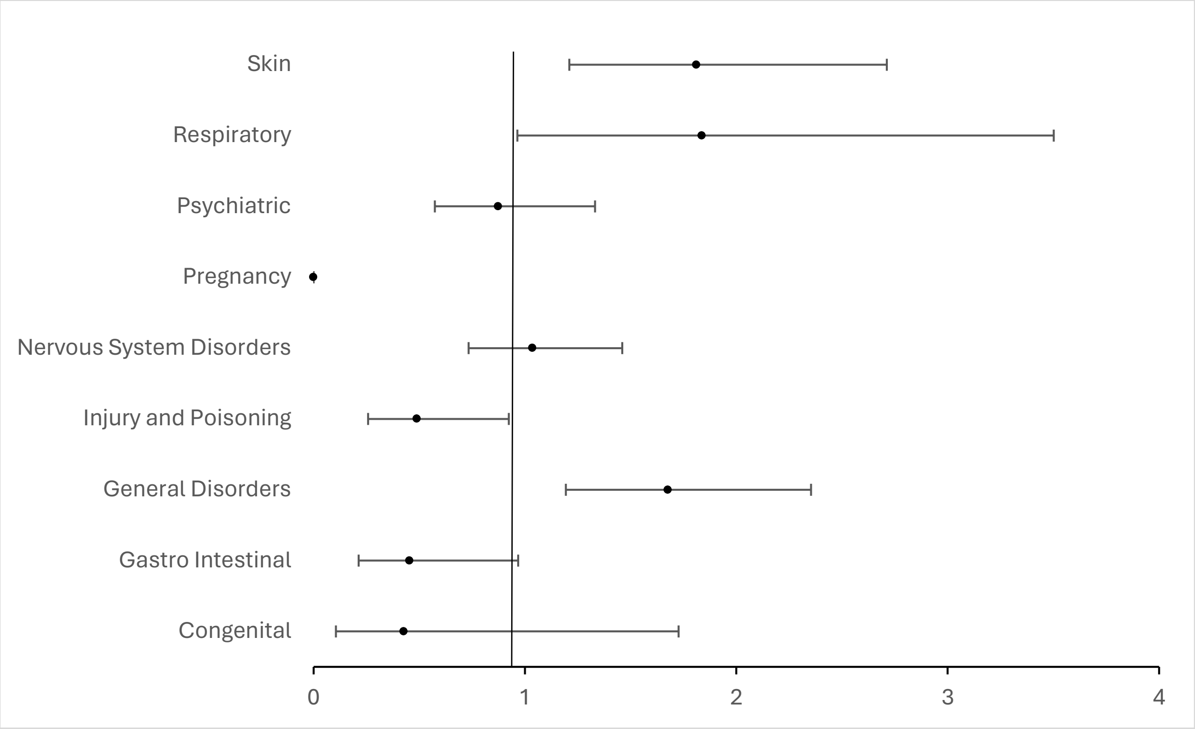

**Table S4:** The number and type of ADR reports for routes of exposure related to the father (data is from a FOI response from the MHRA regarding foetal exposure to VPA via the father 27/10/2023)

| Route of exposure | Number of UK ADR reports | Type of ADR |
|---|---|---|
| Exposure via father,<br>Exposure via partner,<br>Maternal exposure via father during pregnancy,<br>Paternal drugs affecting foetus,<br>Paternal exposure before pregnancy,<br>Paternal exposure during pregnancy,<br>Paternal exposure timing unspecified,<br>Exposure via semen,<br>Transmission of drug via semen | 5 | Cleft palate<br>“Minor” epilepsy<br>Otospondylomegaepiphyseal dysplasia |
